# Development of the SCI-BodyMap: Measuring Mental Body Representations in Adults with Spinal Cord Injury

**DOI:** 10.64898/2026.01.23.26343891

**Authors:** Sydney Carpentier, Sara Bottale, Nicole Cenci, Mauro Cracchiolo, Daniele De Patre, Julian Pablo Gorosito, Ilaria Grimaldi, Mara Melo, Janna Neher, Bianca Polinelli, Leena Rapacz, Marco Rigoni, Fortunata Romeo, Jacquelyn Sinner, Marina Zernitz, Ann Van de Winckel

## Abstract

There is evidence that adults with spinal cord injury (SCI) have deficits in mental body representations (e.g., altered visuospatial body maps and reduced body awareness), due to the diminished or lack of sensory information reaching the brain. These mental body representation deficits are important and need to be quantified, because they can impact daily functioning and they are associated with neuropathic pain. The currently available evaluation scales measure certain aspects of mental body representations with few having been assessed in adults with spinal cord injury. Furthermore, to our knowledge, no scales have been developed specifically for adults with spinal cord injury. Therefore, to address this gap, we completed two aims. First, we developed a novel evaluation scale (the SCI-BodyMap) to measure SCI-specific deficits in mental body representations. Second, we assessed the psychometric properties of inter-rater reliability, test-retest reliability, concurrent validity, with the Revised Body Awareness Rating Questionnaire, the Multidimensional Assessment of Interoceptive Awareness-2, and the Numeric pain rating scale. We also assessed feasibility, utility, and face validity with the QQ-10. We found good to excellent inter-rater and test-retest reliability, with the exception of two items showing moderate test-retest reliability. We did not find any correlations between the SCI-BodyMap with the Revised Body Awareness Rating Questionnaire or the Multidimensional Assessment of Interoceptive Awareness-2 and found fair correlations between high levels of neuropathic pain on the Numeric pain rating scale with highest level of neuropathic pain on the SCI-BodyMap. The scale proved to have high feasibility, utility, and face validity. Once more psychometric analyses are performed in a bigger sample, the SCI-BodyMap could be recommended for use in research and in the clinic.

## Introduction

Spinal cord injury (SCI) often leads to deficits in sensorimotor function. Due to the loss or partial loss of sensory information reaching the brain, the brain has difficulty creating accurate Mental Body Representations (MBRs).^1–4^ MBRs consist of both body awareness and visual spatial body maps. Body awareness has had many definitions across the field, including the awareness of peripersonal space, informed by external and internal sensations, and how the way the body is positioned.^1,5^ An intact visuospatial body map is crucial because it is responsible for informing the body’s spatial location at each moment in time in order to guide movements, perform actions, and interact with the world.

The deficit in MBRs that adults with SCI experience can influence functioning in daily life, such as the ability to transfer, and has also been found to be associated with the presence of neuropathic pain, which is the most common secondary condition of spinal cord injury, affecting up to 69% of individuals with SCI. Neuropathic pain can be debilitating, excruciating at times, and can be a chronic condition for many.

Clinical trials evaluating body awareness interventions such Cognitive Multisensory Rehabilitation (CMR),^2,6^ have reported weaker neural connections between key brain regions important in the creation of MBRs (i.e., parietal operculum, insula, posterior parietal cortex) at baseline when comparing those neural connections at rest in adults with SCI vs uninjured adults. The researchers also reported strengthening of those brain connections as well as improvements in sensorimotor function and reductions in neuropathic pain after completing the CMR intervention. Similarly, the results of other studies evaluating a mind-body approach, Qigong─known to improve awareness of the body─demonstrated participant-reported reductions in neuropathic pain, and improved function after the 12-week Qigong intervention.^4,7–9^

Given the evidence that MBR deficits exist in adults with SCI with significant impact on recovery and daily life, there is a need to measure the degree of these deficits. Previous research attempted to identify significant difference in MBR between adults with SCI and uninjured adults, as well as identifying changes after Qigong^4,8,9^ or CMR^2,6^ using currently available scales, i.e., the Revised Body Awareness Questionnaire (BARQ-R) and the Multidimensional Assessment of Interoceptive Awareness Version 2 (MAIA-2).^10^ The BARQ-R ^11,12^ was able to capture a baseline difference in scores when compared to uninjured adults, as well a pre-post change after adults with SCI underwent either Qigong or CMR.^13^ However, scores on the MAIA-2, focused on interoceptive awareness, showed a more varied picture: For some domains (e.g., Not-Distracting and Trust) adults with SCI scored lower than uninjured adults, for other domains (e.g., Noticing and Attention Regulation) adults with SCI scored higher, and for yet other domains (e.g., Emotional Awareness) there was no significant difference in scoring between adults with SCI and uninjured adults.

Regarding other available scales measuring aspects of MBR, “the Horizontal Subjective Body Midline and the Umbilicus-Reaching Task (URT)”^14^ aims to access the visuospatial representation of the body in the brain of adults with SCI (n=56). This scale is comprised of three tasks: (1) while lying on a table, participants indicate with a laser pointer on the ceiling where they think their umbilicus would project on the ceiling. (2) The same task performed in the dark. (3) The researcher holds the laser pointer, and participants guide the researcher where the laser point should project to match the location of their umbilicus. The latter is to test their ability to rely solely on their visuospatial map without spatial or proprioceptive cues.

The Body Image Task (BIT), researched in both healthy adults (n=18) and adults with SCI (n=24), instructs participants to view a screen with a fixed drawing of a head and were instructed to treat it as a mirror image of themselves. The participant then estimated their body parts to scale in relation to the head. Results of one study using this task showed that healthy adults overestimated the width of their shoulders and the length of their upper arms,^15^ while in a separate study, adults with SCI perceived their body as elongated relative to width of the body compared to uninjured adults.^16^

The Awareness-Body-Chart ‘ABC’,^17^ tested in a sample of university students, is a measure of body awareness that instructs participants to color in a body region based on whether the participant considers their own body part as something they can “perceive with much detail”, “perceive distinctly”, “perceive”, “perceive indistinctly”, or “cannot perceive”.

Although some scales attempt to measure MBR in uninjured adults and adults with SCI, the currently available options only assess one specific element of MBR deficits, (e.g., the location of the umbilicus)^14–16^ and do not address the SCI-specific experiences. Therefore, there is still a need for an evaluation scale that encompasses various elements of MBR deficits identified in adults with SCI. Once psychometric analyses have been completed, this scale could be used in research and in the clinic to measure baseline deficits of MBRs and changes over time.

Therefore, our aims were to **(Aim 1)** Generate and refine items of the SCI-BodyMap for adults with SCI through iterations based on feedback from physical therapists (PTs) or occupational therapists (OTs) and adults with SCI. **(Aim 2)** Evaluate the psychometric properties of the SCI-BodyMap through interrater reliability, test-retest reliability, concurrent validity, feasibility, utility, and face validity. In addition, we investigated MBR differences between adults with SCI and uninjured adults. Finally, as secondary analysis, we determined sub-group differences in MBR deficits between adults with paraplegia vs tetraplegia, and we correlated the SCI-BodyMap, with other SCI-specific variables.

## Method

### Cross-sectional observational study design

In aim 1, we developed the items of the new SCI-BodyMap using an iterative process where refinement of items was created from stakeholder involvement, i.e., PT/OTs and adults with SCI, both through feedback via remotely-delivered surveys and in-person feedback.

In aim 2, we assessed inter-rater reliability for the in-person sections of the scale and test-retest reliability for the self-report sections. We assessed concurrent validity of the *Body awareness items* of the SCI-BodyMap with the BARQ-R,^11,18^ and the MAIA-2,^19,20^ and the *Pain items* of the SCI-BodyMap with the Numeric Pain Rating Scale (NPRS).^21^ The QQ-10 was used to assess feasibility, utility, and face validity.^22^ We compared the SCI-BodyMap scores of adults with SCI to those in uninjured adults. As secondary analyses, we investigated differences in MBR deficits, assessed with the SCI-BodyMap, between adults with paraplegia and tetraplegia, and associations between the SCI-BodyMap and functional mobility, minutes of physical activity per week, and highest level of neuropathic pain. Further details about the study methods can be found in the published protocol found in Carpentier et al. (2025).^18^

### Study Setting

Remote data collection: Surveys to collect demographic, clinical, and SCI-specific information (aims 1 and 2) and item generation feedback (aim 2) were conducted via Zoom.

In-person testing of the SCI-BodyMap (aims 1 and 2): This was conducted in the Brain Body Mind Laboratory at the University of Minnesota-Twin Cities in Minneapolis, Minnesota.

### Recruitment and inclusion/exclusion criteria

*Aim 1:* Through collaborations with the Brain Body Mind Laboratory and the MN Regional Model System (MN Regional SCIMS), we recruited 8 Board-Certified Neurologic Clinical PT/OT. *Inclusion criteria*: PT/OT with clinical experience working with SCI and/or specific experience with CMR, and who had access to internet/iPad/computer/phone.

*Aim 1 and Aim 2:* We recruited 8 adults with SCI for aim 1 and 32 adults with SCI for aim 2 primarily from or contact list or they contacted us through clinicaltrials.gov. *Inclusion criteria*: Adults (18+) with an incomplete or complete SCI of ≥ 1 year who were medically stable, able to read and understand English, and willing to come in-person for testing at the University of Minnesota (for those in aim 2). *Exclusion criteria*: Any medical complications preventing them from sitting for prolonged periods of time or using a ventilator.

*Aim 2:* We also recruited 32 uninjured adults from our contact list of volunteers or they contacted us through study fliers, or the University of Minnesota’s StudyFinder. *Inclusion criteria:* Adults (18+) able to read and understand English. *Exclusion criteria:* Any major medical complications or neurological disorders.

### Demographic and Clinical Outcomes

*For Aims 1 and 2:* For all participants, we collected demographic information including age, sex, gender identity, general health information, including physical activity (min/week), whether or not they practice breathing exercises and whether or not they currently practice body awareness training or have in the past training (e.g., Yoga, Tai-Chi, Qigong, meditation, etc.).

*Aim 1:* Adults with SCI also provided SCI-specific information including years since injury, level of injury, and type of injury (i.e., tetraplegia, paraplegia, Brown-Sequard syndrome, central cord syndrome) and a self-report of the International Standards for Neurological Classification of Spinal Cord Injury (ISNCSCI) ASIA Impairment Scale (AIS) level.^23^

Participants provided information on levels of neuropathic pain, assessed with NPRS,^24^ which is a 3-item self-report measure that assesses lowest, average, and highest neuropathic pain (ranging from 0: no pain to 10: worst pain imaginable) reported in the last week. We assessed participant’s functional mobility using the National Institute of Neurological Disorders and Stroke Common Data Elements (NINDS-CDE) Spinal Cord Injury Functional Index for samples using Assistive Technology (SCI-FI/AT),^25^ which is a self-report measure with 4 domains: Basic Mobility, Self-care, Fine Motor Function, Motor Function, and Ambulation (ranging from 0: unable to do to 4: without any difficulty). There are separate versions for paraplegia and tetraplegia, with content of items altered slightly between the two.

PT/OTs provided demographic outcomes and information about their professional experience (e.g., their professional title, credentials, place of work, and number of years worked with adults with SCI and/or CMR, and an open question for any other relevant information they may want to include).

*Aim 2:* In addition to SCI information collected in Aim 1, all participants completed the **Multidimensional Assessment Interoceptive Awareness-2 (MAIA-2)**,^19^ which is a 37-item self-report measure of interoceptive awareness (ranging from 0: never to 5: always). It contains 8 domains, each measuring a different aspect of interoceptive awareness, including Noticing, defined as the perception of inner bodily cues; Not-Distracting, referring to the capacity to stay engaged with uncomfortable sensations rather than disengage; and Not-Worrying, which reflects reduced emotional sensitivity to unpleasant sensations. Attention Regulation represents the skill in directing and sustaining attention toward bodily cues, while Emotional Awareness involves understanding how physical sensations relate to emotional states. Self-Regulation describes the use of bodily cues to modulate stress and arousal, and Body Listening pertains to interpreting and acting on bodily signals. Finally, Trust reflects a sense of feeling grounded and secure within one’s own body.^19^

Participants also completed the **Revised Body Awareness Rating Questionnaire (BARQ-R)**,^11,12^ which is a 10-item self-report measure of bodily awareness measured on a 4-point Likert scale (ranging from 0: completely disagree to 3: completely agree). Items of this scale consist of questions regarding, bodily tension, digestion, relaxation, body predictability, and awareness of movement and breathing.^11,12^

### Data collection and statistical analyses: Aim 1

#### Item Generation

The resulting evaluation scale is a multidimensional scale composed of two components: an in-person assessment administered by a PT/OT and SC as a trained researcher). The participant also completed the self-report section. The in-person assessment scored by clinicians/researchers includes four domains *[Domain 1: Body Dimension (Items 1a-1h), Domain 2: Body Awareness and Bodily Spatial Relations (Items 2a-2f), Domain 3: Spatial Awareness (item 3), Domain 4: Body Localization (items 4a-4b)]* and the self-report section completed by participants includes three domains *[Domain 5: Body awareness (item 5), Domain 6: Sensation (item 6), and Domain 7: Neuropathic pain (items 7a-7i)*].

We reported the items in the SCI-BodyMap and then, for each iteration, we reported a descriptive summary of changes, highlighting the most notable in the text of this manuscript.

### Data collection and statistical analyses: Aim 2

#### Inter-rater reliability testing

Two raters, SC and one of three authors (LR, JS, JN) assessed 32 adults with SCI with the SCI-BodyMap at the same time.

*Statistical analyses:* We reported Intraclass Correlation Coefficient (ICC) values for items with ratio data (Domain 1). For Domains with ordinal scores (Domains 2-4), we reported Cohen’s kappa coefficient values for items within each Domain. We reported Intraclass Correlation Coefficient (ICC) for total scores of each Domain.

### Test-retest reliability

Adults with SCI completed the self-report items (Domains 5-7) of the SCI-BodyMap on the same day as the in-person evaluation and then at a second time-point at least 1 week later.

*Statistical analyses:* ICC was used to calculate test-retest reliability.

### Concurrent validity

Participants completed the SCI-BodyMap alongside the MAIA-2,^19,20^ and the BARQ-R.^11,12^ Participants also completed the NPRS to report lowest, average, and highest pain the last week.

*Statistical analyses:* We reported Spearman rho correlations for each dimension of the MAIA-2 and the total score of the BARQ-R to assess correlations between the two scales and the SCI-BodyMap in-person items and the Body awareness self-report item. We also reported Spearman rho correlations between the NPRS lowest, average, and highest level of pain separately with the Neuropathic Pain items in the SCI-BodyMap.

### Feasibility, utility, and face validity analyses

The QQ-10 is a measurement scale that assessed the feasibility, utility, and face validity of the SCI-BodyMap.^22^ This questionnaire consists of 10 items on a Likert scale ranging from strongly disagree to strongly agree as well as three open ended questions. Questions asked about relevancy, ease of use, comfortableness, and efficiency of the SCI-BodyMap.

*Statistical analyses:* The QQ-10 generated a value score and a burden score. Each response was coded (strongly disagree=0, mostly disagree=1, neither agree or disagree=2, mostly agree=3, and strongly agree=4). To generate the value score, we took a sum of items 1-6 and to generate the burden score, we took a sum of items 7-10. We then divided each sum by the total available score for the value and burden, respectively, to create a percentage. Greater feasibility, utility, and face validity are reflected by value scores closer to 100 and burden scores closer to 0. In addition to these quantitative ratings, we presented responses to the open-ended questions as quotes.

### Comparison adults with SCI vs healthy adults

We used Mann-Whitney U test to identify differences in scores on the SCI-BodyMap between adults with SCI and healthy adults.

### Secondary analysis: subgroup adults with SCI + correlations with other outcomes

We used a Mann-Whitney U test to identify whether people with paraplegia score differently on the SCI-Body map compared to those with tetraplegia.

Furthermore, we used Spearman rho correlations to investigate associations between the SCI-BodyMap and highest level of neuropathic pain, functional mobility level, minutes of physical activity per week.

## Results

### AIM 1

Completing both components of the SCI-BodyMap required approximately 1–1.5 hours. Each domain is scored separately, noting that each item within Domain 7 is also scored separately.

### Iterative process during scale development

*Iteration 1 (Preliminary data)*: A physical therapist and CMR-certified therapist (SB) outlined initial items based on clinical experience with providing CMR interventions to patients.

*Iteration 2 (Preliminary data)*: The items developed by SB were then sent to a larger group of seven PT from Italy, (BP, DDP, FR, IG, MR, MZ, and NC) and three PT from Brazil (MC, MM, and JG), who were all CMR-certified and who had clinical experience in working with adults with SCI. During this iteration, feedback was mostly content related. For example, in iteration 1 of the scale, item 1 had only 4 body regions, but after implementing suggestions to divide certain areas, iteration 2 had 8 body regions (neck/head, trunk, pelvis, upper arm, forearm, upper leg, lower leg, and foot). Another change was that the self-report section only had one item in iteration 1, but after implementing feedback, we added 2 items about neuropathic pain and sensation, respectively, given its association with MBR and relevance for many adults with SCI. SC created a summary of changes and presented the new iteration back to the Italian CMR therapists, the Brazilian therapists, and SB. All parties agreed to the changes.

*Iteration 3a (Aim 1)*: The resulting items from iteration 2 were then sent to a total of eight clinicians, including seven PTs and one OT. Participants were, on average, 38±8 years old and included six non-Hispanic White females and one female who identified as “other” race (who declined to specify) All clinicians held a Doctor of Physical Therapy (DPT) degree and had an average of 8±5 years of experience working with adults with SCI. Three clinicians were employed in outpatient settings, four in inpatient rehabilitation, and one practiced in both settings. Exposure to CMR varied across participants: four clinicians reported no prior knowledge of CMR, one clinician had been introduced to the approach through a lecture, two had completed a 6-day basic course training, and one clinician had received one month of immersive CMR training in Italy.

Additionally, the self-report items were distributed to five adults with SCI who had previously received CMR and to three adults with no prior CMR exposure. The sample included four men and four women (seven non-Hispanic White participants and one Hispanic woman who identified “Hispanic” as her race), with a mean age of 53±13 years and an average time of 12±9 years post-SCI. Four participants had paraplegia and four had tetraplegia. The injury levels were cervical (n=4), thoracic (n=2), and lumbar (n=2). Most of the feedback given by PT/OTs was about clarity of instructions. Adults with SCI gave feedback on the content of the scale with some indicating that the scale was relevant to them and two people indicating that alternative statements would make things easier to understand. For example, we received feedback that the word “altered” when referring to body awareness did not make sense, so we updated the statement to be more specific about what “altered” could be, e.g., feeling “disconnected” as one participant suggested. We also received feedback from an adult with SCI that the self-report section should have 3 separate items (body awareness, sensation, and neuropathic pain), rather than a combined version of pain and sensation, and therefore, we implemented that suggestion. One PT suggested using a numerical scale rather than coloring, however, one PT and one adult with SCI explicitly stated that they liked the coloring aspect, so we kept those items. Lastly, a participant with SCI suggested having objective descriptions of pain like frequency and duration rather than a notes section, so we added those in as well.

*Iteration 3b (Aim 1):* A subgroup of three adults with SCI and three PTs who gave feedback in iteration 3a came in person to respectively receive or administer the SCI-BodyMap to further improve items and receive feedback. Based on the in-person feedback, we refined instructions and re-ordered tasks for efficiency and ease. We also created printed images to present to participants to help make the instructions clearer for each task. Most notably, we removed one task that involved the PT placing their hand at a distance in front of the participant. The participant was instructed to indicate if the hand was in front of their body or off to the side. This task was found to be too difficult for the PTs to administer due to high rates of error, so we removed it. We also updated one of the tasks (2c) in Domain 2 to avoid redundancy (e.g., an earlier iteration asks the participant to determine which foot was in front of the other foot, which was very similar to item 2a, which asks the participant to determine which foot is closer to an external object placed in front of their body). Both of these tasks required the participant to indicate which foot is either in front or behind the other, so we revised it to now require a participant to determine where their foot is in relation to a pen that sits beneath their foot, more so focusing on the mental body map of the bottom of the foot in relation to a pen rather than the awareness of which foot or leg is ahead of the other. Additionally, participants noted that images of feet and hands with labels describing the task would be helpful, so we printed some out to have on hand and added these to the instruction sheet for the PT/OT to hand out when they administer the evaluation.

### Items of the SCI-BodyMap

Please find the complete SCI-BodyMap with administration and scoring instructions in **Supplementary material.** Below are summarized explanations of each item.

### In-person assessment section

*<u>Domain 1: Body Dimension (Items 1a-1h).</u>* These items reflect a participant’s ability to estimate the dimension of the body. Participants were instructed to imagine, without doing the movement, how many of their own hands it would take to measure each of the 8 body regions (head/neck, trunk, pelvis, upper leg, lower leg, upper arm, forearm, foot). This requires the brain to have an accurate representation of the size, shape, and length of each body region tested as well as their hand.

*Domain 2: Body Awareness and Bodily Spatial Relations (Items 2a-2f)*: Items 2a-2c assess lower limbs and items 2d-2f assess the upper limbs.

2a and 2d: The assessor will place an object on the floor in front of the participant. With eyes closed the participant must determine which foot (or hand) is closer. 2b and 2e: With eyes closed, the participant must determine which part of one foot (or hand) the other foot (or hand) aligned with (e.g., “Are the toes of one foot aligned with the toes, ball, arch, ankle, or heel of your other foot?”).

2c and 2f: The assessor will hold the participant’s foot (or hand) in the air and place a pen directly under the arch (or palm). With eyes closed, the assessor will move the participant’s foot (or hand) over the pen. Once the movement is complete, the participant must determine which part of their foot (or hand) is now hovering over the pen (e.g., “If I were to lower your foot down, do you think the pen is now in front of the toes, at the ball of your foot, the arch, or the heel?”).

To answer correctly, these tasks require that the participant has an accurate MBR of both feet and both hands. They must also have an awareness of their body movements and feelings. For example, when asked informally, what strategy they use to get to their answer, they may place attention to which degree one leg is “stretched” away from the body compared to the other, or how wide of an angle their knee sits at.

*<u>Domain 3: Spatial Awareness (Item 3):</u>* First, the assessor gently guides the participant’s own hand, above different parts of their lower body. The participant, with eyes closed, must determine where their own hand is hovering. (e.g., “If I were to lower your hand, would it miss your body, or would it touch your body, and if the latter, where?”).

To answer correctly, the participant must know the dimension and position of where their lower limbs are sitting in space as well as be able to correctly recognize the orientation of their own hand in relation to the rest of their body.

*<u>Domain 4: Body Localization Task (Items 4a-4b):</u>* First, the assessor will create a snapping sound at either head level or foot level and in either the front of the body or behind the body (4a) or will stay on one side of the body to create a snapping sound at either head, hip, or foot level (4b). With eyes closed, the participant must determine the correct location.

To answer correctly, the participant must have an awareness of where their body is in space and in relation to an external auditory stimulus.

### Self-report assessment section

*<u>Domain 5: Body Awareness (Item 5):</u>* Participants were instructed to shade in areas of a blank body based on the statement about body awareness most applies (e.g., uncolored: *“Even when I am not looking directly at this part of the body, I can clearly feel the shape, size, and dimension and I feel confident of where my body is in space”*; Yellow: *“When I am not looking directly at this part of the body, it feels either blurry, non-defined, or altered in shape, size, or dimension*.”; Blue: “*When I am not looking directly at this part of the body, I feel disconnected to that body part or it feels like it doesn’t exist*.”).

*<u>Domain 6: Sensation (Item 6):</u>* Participants were instructed to shade in areas of a blank body based on the statement about sensation (touch and pressure) that most applies (e.g., uncolored: *“I have full sensation in this part of the body.”;* Yellow: *“I have reduced sensation in this part of the body (partial numbness)”*; Blue: *“I cannot feel any touch or pressure in this part of the body (completely numb).*”).

*<u>Domain 7: Neuropathic Pain (Items 7a-7i)</u>*. Participants were instructed to shade in areas of a blank body based on the statement about neuropathic pain that most applies (e.g., uncolored: “*I have no neuropathic pain in this part of the body.”;* Yellow: *“I feel tingling or non-painful pins and needles in this part of the body (could be perceived as either annoying or just present)**”**;* Blue: *“I have neuropathic pain in this part of the body (for example, burning, stabbing, painful pins and needles, electric shocks.”*) (7a).

The following sections will be completed for both “yellow” pain (7b-e) and “blue” pain (7f-i) separately.

7b and 7f: Assessment of the frequency of neuropathic pain in a typical week for each pain area shaded (e.g., “*had no neuropathic pain”; “less than once a day”;1-5 times a day; or “5 or more times a day”;* or *“100% of the time*).

7c and 7g: Assessment of duration of each pain episode for each pain area shaded (e.g., “*had no neuropathic pain”*; *“less than 1 hour”*; *“1-5 hours”*; *“over 5 hours-almost always”*; *“100 percent of the time*”).

7d/7h and 7e/7i: Assessment of neuropathic pain intensity experienced in a typical week most often and the highest, respectively, for each pain area shaded (e.g., “*had no neuropathic pain*”; “*light*; “*moderate*”; “*severe*”; “*excruciating*”).

### AIM 2

#### Demographic and clinical data

There were 32 adults with SCI who were, on average, 51±15 years old, 37% female, and 15±13 years post-injury. 94% reported neuropathic pain, 66% had an incomplete injury, whereas 34% had a complete injury. There were also 32 uninjured adults who were, on average, 50±20 years old, with 63% identifying as female. **Table 1** displays additional demographic, clinical, and behavioral characteristics of adults with SCI and uninjured adults included in aim 2 of this study.

**Table 1.**
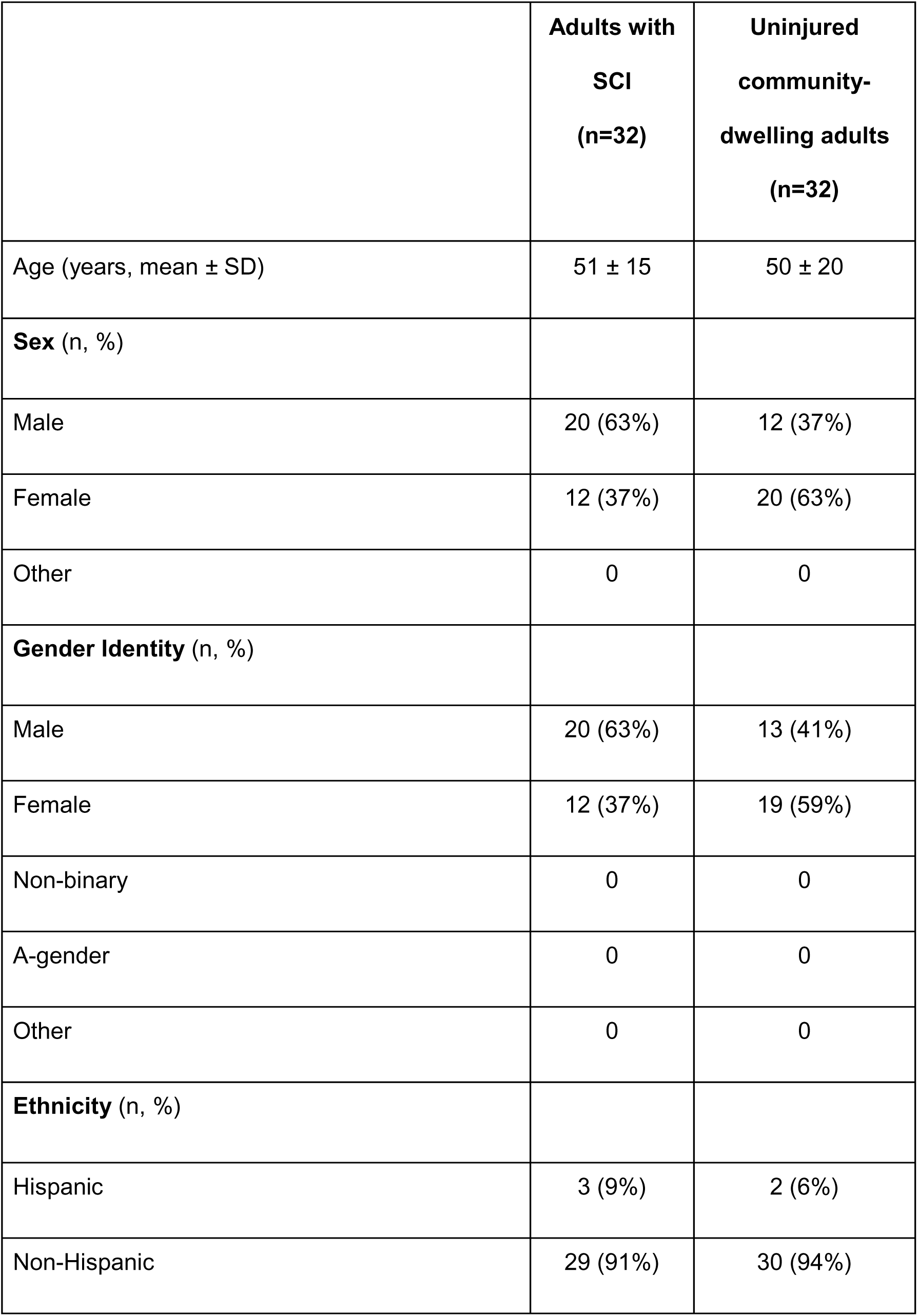
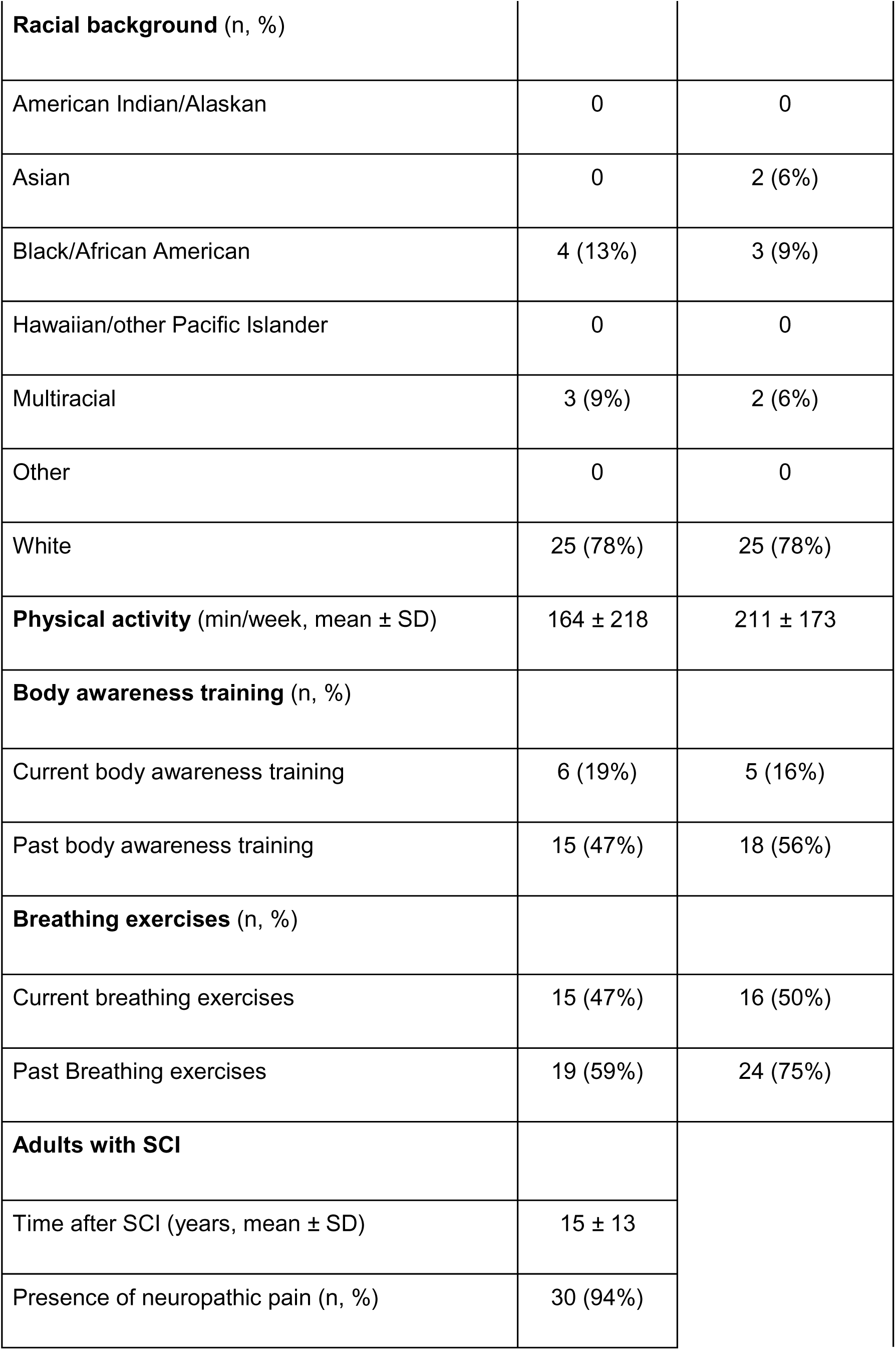
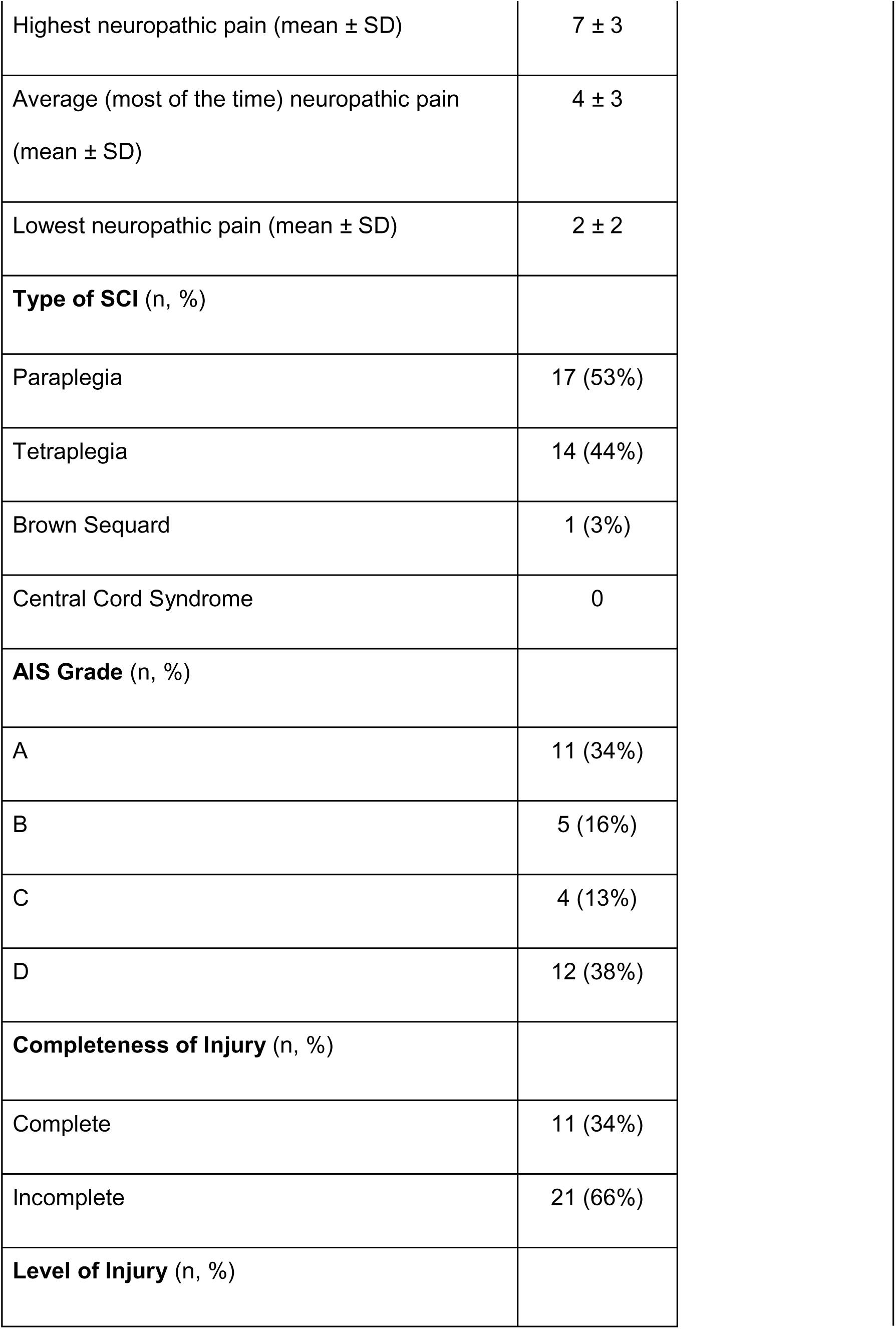
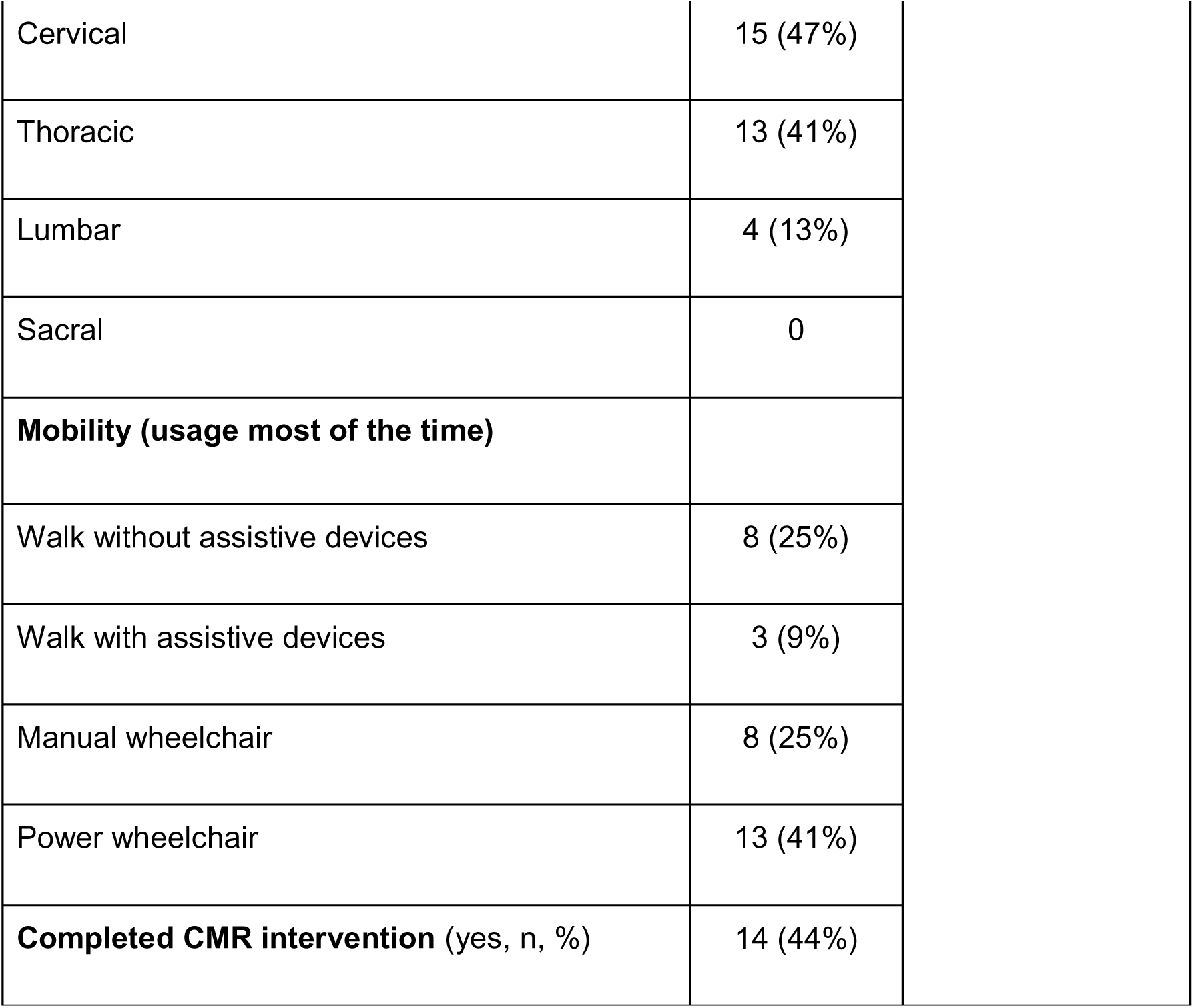
Demographic and clinical information of adults with SCI and uninjured adults.

|  | **Adults with**<br>**SCI**<br>**(n=32)** | **Uninjured**<br>**community-**<br>**dwelling adults**<br>**(n=32)** |
| --- | --- | --- |
| Age (years, mean ± SD) | 51 ± 15 | 50 ± 20 |
| **Sex (n, %)** |  |  |
| Male | 20 (63%) | 12 (37%) |
| Female | 12 (37%) | 20 (63%) |
| Other | 0 | 0 |
| **Gender Identity (n, %)** |  |  |
| Male | 20 (63%) | 13 (41%) |
| Female | 12 (37%) | 19 (59%) |
| Non-binary | 0 | 0 |
| A-gender | 0 | 0 |
| Other | 0 | 0 |
| **Ethnicity (n, %)** |  |  |
| Hispanic | 3 (9%) | 2 (6%) |
| Non-Hispanic | 29 (91%) | 30 (94%) |
| **Racial background** (n, %) |  |  |
| American Indian/Alaskan | 0 | 0 |
| Asian | 0 | 2 (6%) |
| Black/African American | 4 (13%) | 3 (9%) |
| Hawaiian/other Pacific Islander | 0 | 0 |
| Multiracial | 3 (9%) | 2 (6%) |
| Other | 0 | 0 |
| White | 25 (78%) | 25 (78%) |
| **Physical activity** (min/week, mean ± SD) | 164 ± 218 | 211 ± 173 |
| **Body awareness training** (n, %) |  |  |
| Current body awareness training | 6 (19%) | 5 (16%) |
| Past body awareness training | 15 (47%) | 18 (56%) |
| **Breathing exercises** (n, %) |  |  |
| Current breathing exercises | 15 (47%) | 16 (50%) |
| Past Breathing exercises | 19 (59%) | 24 (75%) |
| **Adults with SCI** |  |  |
| Time after SCI (years, mean ± SD) | 15 ± 13 | |
| Presence of neuropathic pain (n, %) | 30 (94%) |  |

|  |  |
| --- | --- |
| Highest neuropathic pain (mean ± SD) | 7 ± 3 |
| Average (most of the time) neuropathic pain<br>(mean ± SD) | 4 ± 3 |
| Lowest neuropathic pain (mean ± SD) | 2 ± 2 |
| **Type of SCI** (n, %) |  |
| Paraplegia | 17 (53%) |
| Tetraplegia | 14 (44%) |
| Brown Sequard | 1 (3%) |
| Central Cord Syndrome | 0 |
| **AIS Grade** (n, %) |  |
| A | 11 (34%) |
| B | 5 (16%) |
| C | 4 (13%) |
| D | 12 (38%) |
| **Completeness of Injury** (n, %) |  |
| Complete | 11 (34%) |
| Incomplete | 21 (66%) |
| **Level of Injury** (n, %) |  |

|  |  |
| --- | --- |
| Cervical | 15 (47%) |
| Thoracic | 13 (41%) |
| Lumbar | 4 (13%) |
| Sacral | 0 |
| **Mobility (usage most of the time)** |  |
| Walk without assistive devices | 8 (25%) |
| Walk with assistive devices | 3 (9%) |
| Manual wheelchair | 8 (25%) |
| Power wheelchair | 13 (41%) |
| **Completed CMR intervention (yes, n, %)** | 14 (44%) |

#### Inter-rater reliability

Domains 1-4, reflecting scoring by clinicians/researchers, had good to excellent inter-rater reliability (ICC=0.84-0.99, **Table 2**), and excellent to perfect agreement (κ=0.84-1.00) of items within Domain 2-4 **(Table 3).**

**Table 2.** Inter-rater reliability (ICC) values of the body awareness items of the SCI-BodyMap (Domains 1-4)

| **SCI-BodyMap Domain** | **ICC** |
| --- | --- |
| *Domain 1: Body Dimension* |  |
| a. trunk | 0.97 |
| b. neck/head | 0.84 |
| c. upper leg | 0.97 |
| d. lower leg | 0.97 |
| e. pelvis | 0.89 |
| f. upper arm | 0.94 |
| g. forearm | 0.95 |
| h. foot | 0.94 |
| *Domain 2: Body Awareness and Bodily Spatial Relations* |  |
| a-c. lower body | 0.99 |
| b-f. upper body | 0.92 |
| *Domain 3: Spatial Awareness* | 0.88 |
| *Domain 4: Body Localization Task* | 0.99 |

**Table 3.** Inter-rater reliability (Kappa values) of the body awareness items of the SCI-BodyMap (Domains 2-4)

| **SCI-BodyMap Domain** | **Kappa (95%CI)** |
| --- | --- |
| *Domain 2: Body Awareness and Bodily Spatial Relations (lower body)* |  |
| Item a. | 0.96 [0.81-1.00] |
| Item b. | 1.00 [0.88-1.00] |
| Item c. | 1.00 [0.87-1.00] |
| *Domain 2: Body Awareness and Bodily Spatial Relations (upper body)* |  |
| Item d. | 0.81 [0.47-1.00] |
| Item e. | 0.84 [0.57-1.00] |
| Item f. | 0.85 [0.43-1.00] |
| *Domain 3: Spatial Awareness* | 0.79 [0.52-1.00] |
| *Domain 4: Body Localization Task* |  |
| Item a. | 0.96 [0.81-1.00] |
| Item b. | 1.00 [0.79-1.00] |

#### Test-retest reliability

For test-retest reliability, two items had moderate test-retest reliability, namely the coloring of normal/abnormal levels of body awareness (Domain 5) and the neuropathic pain frequency (Domain 7, item f). All other items of the self-report (Domains 5-7) had good to excellent test-retest reliability, with ICC ranging from 0.87-0.99 **(Table 4).**

**Table 4.** Test-retest reliability of self-report items of the SCI-BodyMap (Domains 5-7)

| **Domain** | **ICC** |
| --- | --- |
| Domain 5. BA coloring | 0.67 |
| Domain 6. Sensation coloring | 0.91 |
| *Domain 7. Neuropathic Pain (NP)* |  |
| Item 7a. NP Blue and Yellow Coloring | 0.94 |
| Item 7b. (yellow) NP pain frequency | 0.99 |
| Item 7c. (yellow) NP pain duration | 0.99 |
| Item 7d. (yellow) NP level occurring most often | 0.91 |
| Item 7e. (yellow) NP highest level | 0.90 |
| Item 7f. (blue) NP pain frequency | 0.74 |
| Item 7g. (blue) NP pain duration | 0.91 |
| Item 7h. (blue) NP level occurring most often | 0.78 |
| Item 7i. (blue) NP highest level | 0.83 |

**Legend:** BA: body awareness, NP: Neuropathic pain; SCI: Spinal Cord Injury

#### Concurrent validity

After adjusting the alpha level to .002 using the Bonferroni correction, we found no correlations between the MAIA-2 and the BARQ-R with the items on the SCI-BodyMap.

Furthermore, using α=0.05, we found fair correlations between highest pain intensity on the NPRS and the highest neuropathic pain intensity on the SCI-BodyMap (ρ=0.40, *p*=0.023) and no correlation between average pain on the NPRS with pain occurring most often on the SCI-BodyMap (ρ=0.32, *p*=0.079).

#### Feasibility, utility, and face validity of the SCI-BodyMap

The SCI-BodyMap evaluation by participants with SCI on the QQ-10 has a high value score of 75% and a very low burden score of 2.96%, indicating that feasibility, utility and face validity was met.^22,26^ The comprehensive list of feedback given during the QQ-10 is listed in the **Table 5**. Most feedback was given verbally. SC wrote it in 3^rd^ person but did not alter the meaning and provided direct quotes when able.

**Table 5.** Complete list of comments from QQ-10 questionnaire.

| Participant | Do you have any comments or suggestions on how the questionnaire you used could be improved (e.g., structure, appearance or design)? Do you have any other comments or general feedback? | Were any of your important symptoms, problems, or concerns missed out by the questionnaire you used? | Do you feel that any areas or problems in the questionnaire you used were over-represented? |
| --- | --- | --- | --- |
| 1 | For the Body awareness shading statements: "Add one in between yellow and blue, perhaps. He says his feet feel so numb that they aren't disconnected, but he is not engaged with them. I am not afraid to walk but have to be very aware of where I am stepping." He says that this may be automatic for people without SCI, but he has to be really cautious on where he steps. | NA | NA |
| 2 | "Thought it was interesting. Some of the [in-person] items | "proprioception and balance is missing | NA |
|  | [I] had no idea what the answer was. Important for SCI, PT, and recovery, thanks for doing this." She also noted that the part of her body she struggles with is feedback (sensation) that she is either not getting correctly or at all from her feet/legs to the rest of her body/brain. | and is something that I am working on" |  |
| 3 | "A good reminder of how my condition is" | NA | NA |
| 4 | "Sometimes [my] pain is less than 5 seconds and sometimes it is over hours long." | NA | NA |
| 5 | "The snapping part is a big positive because people like to get out and having to be aware of surroundings is important, like needing to know when to back up if someone is behind you or if there are cars." | we "did everything" | NA |
| 6 | NA | NA | NA |
| 7 | "Everyone should have this done once a year." | NA | NA |
| 8 | "It is good to know people are looking at my injury from a different frame of mind." | NA | NA |
| 9 | "I don't feel strongly enough to put strongly agree/disagree for questionnaires." | NA | NA |
| 10 | Requests an "option to split in two days if it becomes too long." | NA | NA |
| 11 | He thought it was pretty extensive and good that pain was broken down to duration and severity for pain. It opened his eyes to other things that were impacted by their injury. "It took a lot of concentration to complete."<br>He noted that he realized he wasn't as aware of his body as he thought he was and would be willing to complete it even every week as routine care, but if it was any more than<br>that then he would suggest making a "short" version if needed daily. | NA | NA |
| 12 | “The questions were straight to the point.” “For the pain questionnaire, it is good to have the comment box as well to add information about what could be causing the pain (e.g., UTI, weather, etc.).” | NA | NA |
| 13 | “It is cool that you are doing this. Anything that brings more awareness to this is great.” | NA | NA |
| 14 | She thinks the scale helped her to communicate about her condition because she can speak to us and appreciates interactive assessments. She also noted she is not a fan of drug trials or placebos for that reason. She answered that the questionnaire made her "upset" because she is competitive and she said the brain power of wanting to get the answer right is why she answered that way, not because of anything we did. | NA | NA |
| 15 | NA | NA | NA |
| 16 | He says he would complete it as routine care, but probably not weekly. He said he would be fine with [doing the assessment] a few times a year because he feels that he is consistent in his pain and awareness so 18a frequent check in is not necessary for him specifically. | He says there is “sufficient inclusion” of everything he is concerned about. | NA |
| 17 | NA | NA | NA |
| 18 | He “can’t disqualify” anything we asked, and he “knew what we were getting at.” He relayed his frustration with himself for not keeping up with the CMR therapy on his own and said he “knew if he kept up with the CMR on his own [he] may have answered more correctly.” He said that “there<br>was nothing that could have been asked differently, but it is good that we filled it out together so [he] could give us more explanation and the questions were a good starting point for the conversation.” | NA | “Number 2 is not relevant for my upper body.” |
| 19 | They thought it was “thorough and enjoyable” | NA | NA |
| 20 | NA | He said it “didn't include spasticity which is related to [his] neuropathic pain.” |  |
| 21 | “It was easy to understand” | NA | NA |
| 22 | They recommended we make a sound with a dog clicker, instead of snapping with fingers | NA | NA |
| 23 | NA | She said it would have been helpful to include more questions regarding<br>the above head area<br>since she has limited movement in neck<br>which is where her injury occurred. for example, she said, "when bending over and then standing up again I hit my head sometimes thinking I was in a different spot in space." | NA |
| 24 | "They were cool. it was fun. I loved it" | NA | NA |
| 25 | NA | NA | NA |
| 26 | "It seemed fine to me" |  |  |
| 27 | He says it is only "mostly" easy to complete because it is just because it is new but thinks it will get easier over time. He also said "this is a great body scan, haven't thought of this in a while" as he was completing the coloring part. He also noted in | He says it is relevant to his life for transfers and moving in space like where his body is in relation to external objects so making it more functional would make it to 'strongly agree' for whether or | NA |
|  | the NP self-report section,<br>“Why is this tingling here?<br>Maybe we can focus on a PT focused on that. Obviously, there is still some connection there (referring to his back that feels a tingly NP sensation). “Excited about this. You’re on to something” | not the scale includes all aspects of his condition that he is concerned about. |  |
| 28 | “It would be helpful to label the body as interior/exterior, so when shading only the outside of the leg it is easier to explain as well as potentially labeling other anatomical parts on the body. The only embarrassing thing was her having to say "buttocks" when coloring in the drawing. | NA | NA |
| 29 | NA | NA | NA |
| 30 | “I know it’s more invasive, but include bladder, bowel-huge part in SCI recovery.” | NA | NA |
| 31 | NA | NA | NA |
| 32 | NA | “You could add something with<br>moving or standing<br>and balance.” |  |

#### Comparisons of scores of the SCI-BodyMap between adults with SCI vs. healthy adults

Two items on the SCI-BodyMap scored significantly lower in adults with SCI compared to healthy adults. The first item was Domain 2 (lower body), meaning identifying the spatial location of one foot compared to the other foot or to an object, Mann-Whitney *U*=-4.49, *p*<0.0001 with Median (IQR) scores for SCI: 25.5 (12.5) and healthy: 34 (3). The second item was self-report coloring of body awareness (Domain 5), Mann-Whitney *U*=-4.85, *p*<0.0001 with Median (IQR) scores for SCI: 55 (20.25) and healthy: 60 (0).

### Secondary analyses

#### Subgroup adults with SCI

After adjusting the alpha level to .004 using the Bonferroni correction, we found no difference in scores when comparing adults with paraplegia (n=17) vs tetraplegia (n=14 and one person with Brown-Sequard).

#### Correlations with NPRS highest pain, physical activity, and SCI-FI/AT items

After adjusting the alpha level to .003 using the Bonferroni correction, we found the SCI-FI/AT ambulation domain correlated strongly with Domain 2 (lower body), i.e., identifying the spatial location of one foot compared to the other or an object (Spearman ρ=0.71, *p*<0.0001).

## Discussion

The aims of this study were to (aim 1) develop an evaluation scale to measure MBRs in adults with SCI and (aim 2) to assess its inter-rater reliability, test-retest reliability, concurrent validity with the BARQ-R,^11,12^ the MAIA-2,^19,20^ NPRS,^21^ and feasibility, usability, and face validity with the QQ-10.^22^ In addition, we identified whether healthy adults score statistically different on the SCI-BodyMap scale than adults with SCI. As secondary analysis, we identified if there were significant differences in SCI-BodyMap Body Awareness scores in adults with paraplegia vs tetraplegia. Finally, we correlated SCI-BodyMap Body Awareness items to highest level of neuropathic pain, functional mobility level, minutes of physical activity per week.

### Main results

Overall, the scale proved to have good to excellent inter-rater reliability and test-retest reliability, with the exception of two items: coloring normal/abnormal levels of body awareness and neuropathic pain frequency. Neuropathic pain frequency can change week per week due to a variety of factors revealed in the comment section of the SCI-BodyMap (e.g., sleep quality, weather), as confirmed by our participants, and thus this could explain the lower test-retest reliability when tested a week later. Parts of the body that feel normal, blurry, or disconnected can also slightly change over time, depending on factors such as how well a person has slept. Both these items may thus not necessarily reflect a problem with reliability but may possibly reflect some dynamic aspects of pain and body awareness, which can slightly fluctuate week to week. Further research in a larger sample would be necessary to identify whether those items should be taken out, scored differently to capture a more stable reporting, or whether this is just the nature of perception of these pain and body awareness aspects that vary week by week.

The QQ-10^22^ revealed high value and low burden, and the qualitative feedback received supported the face validity, satisfaction and utility of the scale. Adults with SCI viewed the scale as important and relevant to their lived-experience. Although there was generally positive feedback, participant feedback noted that the phrases did not completely describe their experience and suggested adding an additional description, which will be implemented in the next iteration.

With regard to concurrent validity, the lack of correlation between the SCI-BodyMap and the BARQ-R and the MAIA-2 can be explained by the fact that the BARQ-R^11^ and the MAIA-2^19^ measure slightly different aspects of body awareness and interoceptive awareness than the SCI-BodyMap, respectively. The BARQ-R has questions more related to feeling tension in the body, ability to relax, and whether or not the body is predictable^11^ and the MAIA-2 assesses one’s ability to listen to the body, find trust in the body, and self-regulate in times of pain or discomfort.^19^

It was more surprising that the scores for neuropathic pain most often perceived had no correlation to NPRS average pain and that highest neuropathic pain levels in a typical week only fairly correlated with NPRS highest pain intensity levels. Possibly the scoring options for average (most of the time) and highest neuropathic pain may need to be rephrased in the SCI-BodyMap to both either reflect “typical week” or “in the past week” or changed to a 0-10 scoring system like the NPRS.^21^

As expected, adults with SCI had significantly more difficulty identifying the location of the foot in relation to the other foot or to an object when comparing the same task in uninjured adults (e.g., Domain 2, lower body items). This aligns with previous research indicating that it is necessary to have an accurate MBR in order to locate the dimension and location of an external object.^27^

Adults with SCI also scored significantly lower on the self-report measures of coloring the level of awareness of the body (normal, blurry, or disconnected) compared to uninjured adults who scored a maximum on this test. This is also expected, given the MBR deficit in adults with SCI. The responses on this self-report question on the SCI-BodyMap align with previous qualitative research of Vázquez-Fariñas M. and Rodríguez-Martin B. (2021) who conducted qualitative research, finding that adults with SCI feel their body is “fragmented” or “alien,” similar to what we found in assessment of the SCI-BodyMap with adults with SCI who shaded in parts of the body reflecting statements of feeling “blurry” or “disconnected.” ^28^

With regards to secondary analysis of correlations, we found a strong correlation between SCI-FI/AT ambulation and the ability to correctly locate the foot on the floor in relation to the other foot or an object, which is expected, as being able to locate where the foot is on the floor is crucial for walking.

This work has expanded on those of others. Deficits in MBR for adults with SCI has been confirmed by previous research in brain imaging studies,^2^ and in other tasks where participants must relate their body to external stimuli^29^ or to illusory paradigms such the rubber hand illusion.^30–32^ Participants in these tasks combine tactile, and visual information as well as illusory objects to determine deficits in MBR. The SCI-BodyMap is novel in that we assess comparisons within their own body, and participants need to refer to their own body signals and information solve the tasks in the items. It allows for adults with SCI and clinicians to determine quantifiable degrees of MBR deficit and changes over time.

Specifically for our self-report section, there are some evaluation scales that measure aspects of MBR, such as the “Awareness Body Chart (ABC),” however, it was not developed specifically for adults with SCI.^17^ The psychometric properties of this new scale, for example, were validated in physical therapy students who were assumed to be a “homogeneous group” of adults with a “sufficiently good and [a] healthy state of body awareness.” The SCI-BodyMap has similar aspects to this scale in that we have a self-report portion that allows participants to describe how well they are aware of certain body parts. The ABC has statements ranging from “I can perceive with much detail” to “I cannot perceive.” Our SCI-BodyMap expands upon research in that our self-report section consists of statements that are unique to the SCI-experience and is confirmed in our results that those with SCI scored significantly lower than uninjured adults, and that uninjured adults scored on average a maximum score on this item.

Fuentes et al. (2012) used the Body Image Task (BIT)^16^ to assess “bodily distortions” in 24 adults with SCI, given previous knowledge of MBR deficits due to impaired sensorimotor function after SCI. They found that participants reported an overestimation of limb length relative to width compared to healthy controls. However, Fuentes et al. (2013) found that even “healthy” adults struggle to correctly estimate the dimensions of their body parts.^15^ Our findings with the SCI-BodyMap echoed this result when examining the length and dimension of body parts in both adults with SCI and uninjured adults, given that there was no significant difference between the two groups. However, our assessment method differed; participants relied on their kinesthetic sensations of their body and of their imagined length of their hand to identify the length of other body parts. In contrast, the BIT required participants to convert their kinesthetic feeling of their body into a visual image of their body dimensions.

To gain further insight into MBR deficits, future iterations of the SCI-BodyMap could include an assessment of the participant’s awareness of their hand dimensions before using it as a reference for imagining the length of other body parts. This is particularly relevant considering prior research indicates that healthy adults tend to overestimate the dimensions of their hands and length of their fingers.^33^

### Limitations

Because Aim 2 required in-person participation, most participants in our testing were from Minnesota or neighboring states. As a result, both participant input and PT feedback were regionally concentrated and lacked diversity (78% were White non-Hispanic participants). In addition, in Aim 1, the PT group lacked demographic and racial/ethnic diversity as well. A subsequent iteration should therefore incorporate feedback from PTs and adults with SCI from other regions of the United States and would emphasize more recruitment from adults of diverse backgrounds.

## Conclusion

To our knowledge, the SCI-BodyMap is the first scale designed to specifically measure deficits in MBR for adults with SCI. Overall, the SCI-BodyMap demonstrated good to excellent reliability, feasibility, usability, and face validity. The scale was designed by considering international stakeholders, and specifically also input from adults with SCI. An advantage of the SCI-BodyMap is that it was designed to measure multiple different aspects of MBR, encompassing both in an in-person assessment and a self-report assessment. Future iterations of the SCI-BodyMap will incorporate the comments from the QQ-10 evaluation. Further psychometric analyses are needed in a larger, more diverse population, with additional analyses such as minimally detectable change (MDC), or minimal clinically important difference (MCID). In addition, a remote version as well as a short-form of the scale could be designed and evaluated. The SCI-BodyMap could also be translated and evaluated in other languages and for other cultures. Once psychometric analyses have been completed, the SCI-BodyMap could show potential for effective use in the clinic or in research.

## Data Availability

The datasets generated during and analyzed during the current study are available from the corresponding author on request.

## Acknowledgments

We would like to thank all participants and clinicians for their invaluable help. We extend our profound thanks to Marc Noël for the critical review of the manuscript.

## References

1. De Martino ML, De Bartolo M, Leemhuis E, Pazzaglia M. Rebuilding Body–Brain Interaction from the Vagal Network in Spinal Cord Injuries. Brain Sciences. 2021;11(8):1084. doi:10.3390/brainsci11081084

2. Van de Winckel A, Carpentier ST, Deng W, et al. Identifying body awareness-related brain network changes after cognitive multisensory rehabilitation for neuropathic pain relief in adults with spinal cord injury: Delayed treatment arm phase I randomized controlled trial. medRxiv. Published online February 10, 2023. doi:10.1101/2023.02.09.23285713

3. Lucci G, Pazzaglia M. Towards multiple interactions of inner and outer sensations in corporeal awareness. Front Hum Neurosci. 2015;9:163. doi:10.3389/fnhum.2015.00163

4. Van de Winckel A, Carpentier ST, Deng W, et al. Feasibility of using remotely delivered Spring Forest Qigong to reduce neuropathic pain in adults with spinal cord injury: a pilot study. Front Physiol. 2023;14:1222616. doi:10.3389/fphys.2023.1222616

5. Mehling WE, Wrubel J, Daubenmier JJ, et al. Body Awareness: a phenomenological inquiry into the common ground of mind-body therapies. *Philosophy*, Ethics, and Humanities in Medicine. 2011;6(1):6. doi:10.1186/1747-5341-6-6

6. Van de Winckel A, Carpentier S, Deng W, et al. Identifying body awareness-related brain network changes after cognitive multisensory rehabilitation for neuropathic pain relief in adults with spinal cord injury: Protocol of a phase I randomized controlled trial. Top Spinal Cord Inj Rehabil. 2022;28(4):33–43. doi:10.46292/sci22-00006

7. User S. Courses - Five Element Healing Movements. Accessed September 30, 2025. https://legacy.springforestqigong.com/shop/healing-tools-courses/five-element-healing-movements-store

8. Van de Winckel A, Carpentier S, Deng W, Zhang L, Battaglino R, Morse L. Using remotely delivered Spring Forest Qigong^TM^ to reduce neuropathic pain in adults with spinal cord injury: protocol of a quasi-experimental feasibility clinical trial. Pilot Feasibility Stud. 2023;9(1):145. doi:10.1186/s40814-023-01374-3

9. Van de Winckel A, Zhang L, Lim KO, et al. Changing the Perceived Pain Intensity in Populations With Spinal Cord Injury and With Health Disparities (HAPPINESS): Protocol for a feasibility study. JMIR Res Protoc. 2025;14:e82431. doi:10.2196/82431

10. Mehling WE, Price C, Daubenmier JJ, Acree M, Bartmess E, Stewart A. The Multidimensional Assessment of Interoceptive Awareness (MAIA). PLoS One. 2012;7(11):e48230. doi:10.1371/journal.pone.0048230

11. Dragesund T, Strand LI, Grotle M. The Revised Body Awareness Rating Questionnaire: Development Into a Unidimensional Scale Using Rasch Analysis. Phys Ther. 2018;98(2):122–132. doi:10.1093/ptj/pzx111

12. Carpentier S, Deng W, Blackwood J, Van de Winckel A. Rasch validation of the revised body awareness rating questionnaire (BARQ-R) in adults with and without musculoskeletal pain. BMC Musculoskelet Disord. 2024;25(1):799. doi:10.1186/s12891-024-07893-1

13. Carpentier S, Van de Winckel A. Changes in body and interoceptive awareness after Cognitive Multisensory Rehabilitation or Qigong in adults with spinal cord injury. medRxiv. Published online December 22, 2025. doi:10.64898/2025.12.12.25342154

14. Osinski T, Martinez V, Bensmail D, Hatem S, Bouhassira D. Interplay between body schema, visuospatial perception and pain in patients with spinal cord injury. Eur J Pain. 2020;24(7):1400–1410. doi:10.1002/ejp.1600

15. Fuentes CT, Longo MR, Haggard P. Body image distortions in healthy adults. Acta Psychol (Amst). 2013;144(2):344–351. doi:10.1016/j.actpsy.2013.06.012

16. Fuentes CT, Pazzaglia M, Longo MR, Scivoletto G, Haggard P. Body image distortions following spinal cord injury. J Neurol Neurosurg Psychiatry. 2013;84(2):201–207. doi:10.1136/jnnp-2012-304001

17. Danner U, Avian A, Macheiner T, et al. ABC-The Awareness-Body-Chart: A new tool assessing body awareness. PLoS One. 2017;12(10):e0186597. doi:10.1371/journal.pone.0186597

18. Carpentier S, Bottale S, Cenci N, et al. Development of the SCI-BodyMap—measuring mental body representations in adults with spinal cord injury: Protocol for item generation, reliability, and validity testing. JMIR Res Protoc. 2025;14:e72370. doi:10.2196/72370

19. Mehling WE, Acree M, Stewart A, Silas J, Jones A. The Multidimensional Assessment of Interoceptive Awareness, Version 2 (MAIA-2). PLoS One. 2018;13(12):e0208034. doi:10.1371/journal.pone.0208034

20. Blackwood J, Carpentier S, Deng W, Van de Winckel A. Preliminary Rasch analysis of the multidimensional assessment of interoceptive awareness in adults with stroke. PLoS One. 2023;18(6):e0286657. doi:10.1371/journal.pone.0286657

21. Numeric Pain Rating Scale. Shirley Ryan AbilityLab. Accessed December 10, 2024. https://www.sralab.org/rehabilitation-measures/numeric-pain-rating-scale

22. Moores KL, Jones GL, Radley SC. Development of an instrument to measure face validity, feasibility and utility of patient questionnaire use during health care: the QQ-10. Int J Qual Health Care. 2012;24(5):517–524. doi:10.1093/intqhc/mzs051

23. Kirshblum S, Snider B, Rupp R, Read MS, International Standards Committee of ASIA and ISCoS. Updates of the International Standards for Neurologic Classification of Spinal Cord Injury: 2015 and 2019. Phys Med Rehabil Clin N Am. 2020;31(3):319–330. doi:10.1016/j.pmr.2020.03.005

24. Hanley MA, Jensen MP, Ehde DM, et al. Clinically significant change in pain intensity ratings in persons with spinal cord injury or amputation. Clin J Pain. 2006;22(1):25–31. doi:10.1097/01.ajp.0000148628.69627.82

25. Tulsky DS, Kisala PA. Overview of the Spinal Cord Injury-Functional Index (SCI-FI): Structure and Recent Advances. Arch Phys Med Rehabil. 2022;103(2):185–190. doi:10.1016/j.apmr.2021.10.006

26. Twohig H, Jones G, Mackie S, Mallen C, Mitchell C. Assessment of the face validity, feasibility and utility of a patient-completed questionnaire for polymyalgia rheumatica: a postal survey using the QQ-10 questionnaire. Pilot Feasibility Stud. 2018;4:7. doi:10.1186/s40814-017-0150-y

27. Longo MR, Azañón E, Haggard P. More than skin deep: body representation beyond primary somatosensory cortex. Neuropsychologia. 2010;48(3):655–668. doi:10.1016/j.neuropsychologia.2009.08.022

28. Vázquez-Fariñas M, Rodríguez-Martin B. “Living with a fragmented body”: a qualitative study on perceptions about body changes after a spinal cord injury. Spinal Cord. 2021;59(8):855–864. doi:10.1038/s41393-021-00634-4

29. Scandola M, Aglioti SM, Bonente C, Avesani R, Moro V. Spinal cord lesions shrink peripersonal space around the feet, passive mobilization of paraplegic limbs restores it. Sci Rep. 2016;6(1):24126. doi:10.1038/srep24126

30. Giurgola S, Crico C, Farnè A, Bolognini N. The sense of body ownership shapes the visual representation of body size. J Exp Psychol Gen. 2022;151(4):872–884. doi:10.1037/xge0001111

31. Ambron E, Miller A, Connor S, Branch Coslett H. Virtual image of a hand displaced in space influences action performance of the real hand. Sci Rep. 2020;10(1):9515. doi:10.1038/s41598-020-66348-4

32. Lenggenhager B, Scivoletto G, Molinari M, Pazzaglia M. Restoring tactile awareness through the rubber hand illusion in cervical spinal cord injury. Neurorehabil Neural Repair. 2013;27(8):704–708. doi:10.1177/1545968313491009

33. Longo MR, Haggard P. An implicit body representation underlying human position sense. Proc Natl Acad Sci U S A. 2010;107(26):11727–11732. doi:10.1073/pnas.1003483107

