## Supplementary material for "Development of the SCI-BodyMap: Measuring Mental Body Representations in Adults with Spinal Cord Injury"

**SCI-BodyMap**

***Higher score = better***

**Materials to prepare:**

- A (preferably high/low) treatment table
- An easily movable desk or table
- A chair with arm and back support (if needed for item 4 or for those that cannot support themselves on a treatment table)
- An object to use for item 2 (e.g., soda can, pen, or cone)
- A pen to write scores
- A soft tape measure (or other soft string) used to check measurements for item 1(e.g., tape measure, string, exercise band)

**General Instructions for PT:**

- It is important that for each of these items to stay neutral and inviting, whether or not the patient has answered correctly or not.
- Please do not let the patient know whether or not they answered correctly, and do not provide them with a score unless they request at the very end.
- Please do not let the patient have more than one guess or repeat any tasks unless they are asking for clarity of instructions

The script is in Purple. Any other notes are for the PT, but are not meant to be read out loud.

Say the following:

Research has shown that those with SCI experience a deficit in bodily awareness, how the brain maps out the body, and how the body interacts with itself and in space. This is important to note, because we have seen that through therapies that focus on improving body awareness, it in turn has resulted in an improvement in sensory and motor function, as well and in neuropathic pain levels.

In this evaluation, we will have you answer a variety of questions that aim to measure a level of body awareness.

The first part of the evaluation will be administered by a therapist. Then, we will give you a self-report form to complete on your own. Let’s get started with the first part of the evaluation.

### Domain 1: BODY DIMENSION

**Preparation:**

- The patient should lay down supine on a treatment table with a pillow under head and legs for support.
- If patient is unable to self-transfer or it is too difficult, then make a note in the scoring that it was done sitting up in a chair or wheelchair.
- The patient should have their shoes off.

For this first set of questions, I will ask you to imagine how many of *your own* hands it would take to measure certain parts of your body. The goal of this task is to see how you feel or perceive your body, without counting or using other calculations to get to the answer.

Please press all of your fingers together so that there is no space between each finger, including the thumb. You may look at your hand for only a few seconds to get a reference, but please do not spend any more time than that and please do not look at any other parts of your body.

Without actually doing it, imagine how many of your own hands it would take to measure your belly from your hip to your collar bone. I will show you the start and end spot on myself. I will show you as an example on my own body.

*Instructions:* Indicate on your own body where the starting position will be and where the ending position will be. Also, in the air you may place your hands directly next to each other up to mimic the movement they will imagine, but only for one or two hands. Do not allow the participant to look at their own body or hand longer than a couple seconds, to avoid them using calculations to get to their answer. In the case that your participant cannot manually check how many hands by doing the actual movement, or in the case that you would like measurement to be consistent across groups (e.g., in a research setting), use a tape measure to take a measurement of their hand and then of the body part, and divide the number of inches of hand by number of inches of body part to get to the correct answer.

Scoring: For each Item, calculate the number of missed hands.

|  | Starting Positions | Ending Positions | Patient answer | Correct  Answer (measur  e to the nearest half hand) | How many hands off (Score) ? |
| --- | --- | --- | --- | --- | --- |
| a. Trunk | Little finger touching the inguinal area. | Index finger touching collar bone. |  |  |  |
| b. Neck/Head | Little finger touching collar bone. | Hands stops before the curvature of the head. |  |  |  |
| c. Upper Leg | Index finger touching the inguinal area. | Little finger ending above the kneecap. |  |  |  |
| d. Lower Leg | The hand starts  at the kneecap. | Little finger touches ankle where it meets the top of the foot. |  |  |  |
| e. Pelvis | Longest fingertip touching thigh  at Inguinal area. | Hand stopping directly before first  hand would have started. |  |  |  |
| f. Upper  Arm | Little finger at the elbow joint. | Index finger stops at the top of the shoulder joint (deltoid muscle). |  |  |  |
| g. Forearm | Little finger touches the end of the fingertips  of the other  hand. | Little finger at the elbow joint. |  |  |  |
| h. Foot | Bottom of the foot. The hand starts covering the toes perpendicular to the foot. | Hand stopping once heel is covered fully. |  |  |  |

Domain 1: For each Item, calculate the number of missed hands. LOWER score (i.e., less missed hands), is BETTER.

**Domain 2: BODY AWARENESS AND BODILY SPACIAL RELATIONS**

**Preparation:**

- The patient should sit on the edge of the treatment table with feet flat on the floor and legs at least 90-degree angle.
- If the patient is unable to reach the ground, place a box under the feet to ensure the correct positioning.

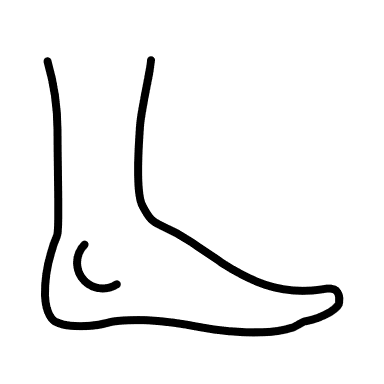

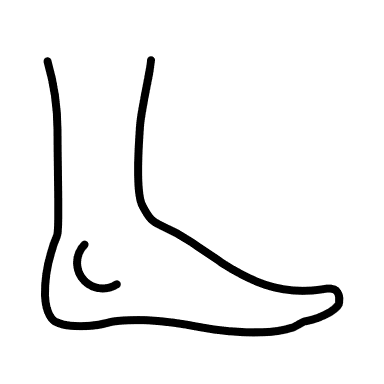

- The patient should have their shoes off.

2ft.

- Place an object about 2 feet away on the floor

**2a.** The goal of this task is to relate one body part to other body parts and understand where the body is in relation to an external object. For this task, when it starts, I will cover your lower body so you are unable to see. For each foot, I can move it either forwards or backwards. Once I have the position, I will ask you which foot is closer to the object or if they are even.

*Instructions:* Do NOT recenter between movements.

| **Movement** | **Score** |
| --- | --- |
| 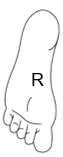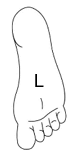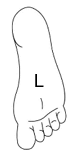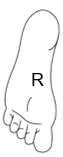 1.  **Left** foot moves and lands behind right foot | 1.Correct answer: R is closer  **Score:**  Correct (1)  Incorrect (0)  **Notes:** |
| 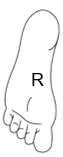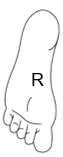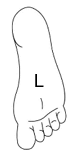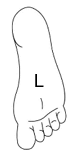2.  **Left** foot moves and lands in front of right foot | 2. Correct answer: L is closer  **Score:**  Correct (1)  Incorrect (0)  **Notes:** |
| 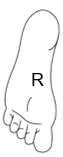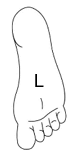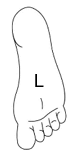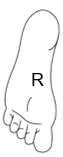3.  **Left** foot moves and lands behind right foot | 3. Correct answer: R is closer  **Score:**  Correct (1)  Incorrect (0)  **Notes:** |
| 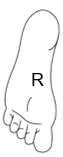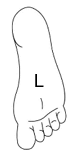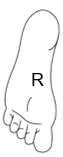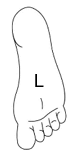4.  **Left** foot moves and lands behind right foot | 4. Correct answer: Even  **Score:**  Correct (1)  Incorrect (0)  **Notes:** |
| 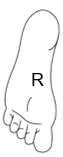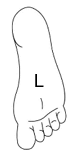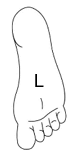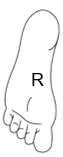5. ***Instructions:* Please re-center before moving to the R foot.**  **Right** foot moves and lands in front left foot | 5. Correct answer: R is closer  **Score:**  Correct (1)  Incorrect (0)  **Notes:** |
| 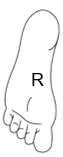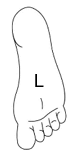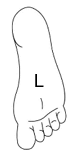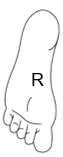6.  **Right** foot moves and lands even with left foot | 6. Correct answer: Even  **Score:**  Correct (1)  Incorrect (0)  **Notes:** |
| 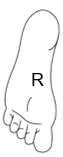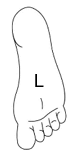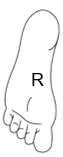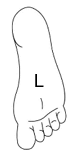7.  **Right** foot moves and lands behind left foot | 7. Correct answer: L is closer  **Score:**  Correct (1)  Incorrect (0)  **Notes:** |
| 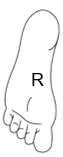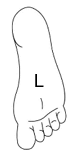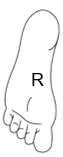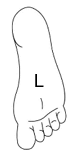8.  **Right** foot moves and lands in front left foot | 8. **Correct answer:** R is closer  **Score:**  Correct (1)  Incorrect (0)  **Notes:** |

**2b**. The goal of this task is to relate one foot to the other. For this task, when it starts, I will keep your lower body covered so you are unable to see. I will move either your right or left foot and then have you determine at what part of one foot, the toes of one foot is touching. Please look at the handout for a visual of your options. The toes of one foot will be in line with either your toes (even), the balls of your foot, the arch of your foot, your ankle, or the back of your foot.

*Instructions:* **Do NOT re-center.**

| **Movement** | **Score** |
| --- | --- |
| 1.Move the **L foot** back and forth to then land at balls of right foot.  The tip of your Left foot, is in line with which part of your right foot?  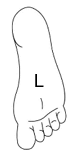  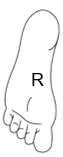  Ball | 1. Correct answer: **balls** of right foot  **Score** (circle what the patient answers):  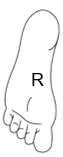  Heel (0)  Ankle (0)  Arch (1)  Ball (2)  Toes (1)  **Notes:** |
| 2.Without re-centering, move the **L foot** back  and forth to then land at arch of right foot.  The tip of your Left foot, is in line with which part of your right foot? 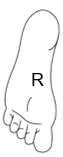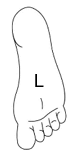 arch | 2. Correct answer: a**rch** of right foot  **Score:**  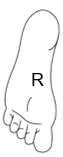  Heel (0)  Ankle (1)  Arch (2)  Ball (1)  Toes (0)  **Notes:** |
| 3.Without re-centering, move the **L foot** back  and forth to then land at the toes of right foot.  The tip of your Left foot, is in line with which part of your right foot? 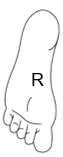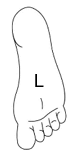 toes | 3. Correct answer: t**oes** of right foot (even)  **Score:**  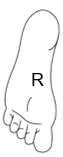  Heel (0)  Ankle (0)  Arch (0)  Ball (1)  Toes (2)  **Notes:** |
| 4.Without re-centering, move the **L foot** back  and forth to then land at the heel of right foot.  The tip of your Left foot, is in line with which part of your right foot? 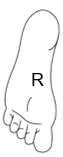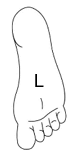 heel | 4. Correct answer: **heel** of right foot  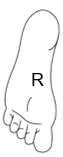  Heel (2)  Ankle (1)  Arch (0)  Ball (0)  Toes (0)  **Notes:** |
| 5. *Instructions:* **Please re-center before moving to the R foot**. Move the R **foot** back and forth to then land at the balls of left foot.  The tip of your Right foot, is in line with which part of your left foot? 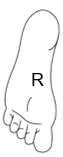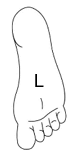 ball | 5. Correct answer: **balls** of left foot  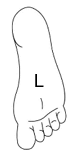  Heel (0)  Ankle (0)  Arch (1)  Ball (2)  Toes (1)  **Notes:** |
| 6.Without re-centering, move the R **foot** back  and forth to then land at the ankle of left foot.  The tip of your Right foot, is in line with which part of your left foot? 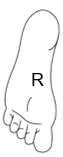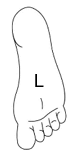 ankle | 6. Correct answer: **ankle** of left foot  **Score:**  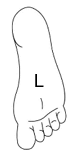  Heel (1)  Ankle (2)  Arch (1)  Ball (0)  Toes (0)  **Notes:** |
| 7.Without re-centering, move the R **foot** back  and forth to then land at the arch of left foot.  The tip of your Right foot, is in line with which part of your left foot? 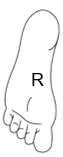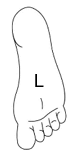 arch | 7. Correct answer: **arch** of left foot  **Score:**  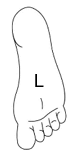  Heels (0)  Ankle (1)  Arch (2)  Ball (1)  Toes (0)  **Notes:** |
| 8.Without re-centering, move the R **foot** back  and forth to then land at the balls of left foot.  The tip of your Right foot, is in line with which part of your left foot? 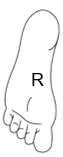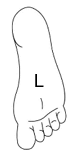 ball | 8. Correct answer: **balls** of left foot  **Score:**    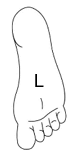Heel (0)  Ankle (0)  Arch (1)  Ball (2)  Toes (1)  **Notes:** |

**2c:** Now I will lift up your foot and place a pen underneath the arch. The pen will remain in the same position, but I will move your foot in the air above it. For each movement, I will ask If I were to set your foot down, which part of your foot would touch the pen. Please use this hand-out for reference.

The options will be in front of toes (not touching), balls, arch, and heel.

| **Movement** | **Score** |
| --- | --- |
| 1.Keep the left foot in place, and move **the R foot** back and forth to then land so the balls of the foot to hoover over the pen.  If I were to set your foot down, which part of your foot would touch the pen?  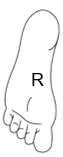 | 1. Correct answer: **ball** of right foot  **Score** (circle what the patient answers):  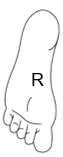  Heel (0)  Arch (1)  Ball (2)  Ahead of toes – not touching (1)  **Notes:** |
| 2.Keep the left foot in place, and move **the R foot** back and forth to then land so the arch of the foot to hoover over the pen.  If I were to set your foot down, which part of your foot would touch the pen?  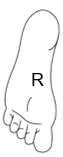 | 2. Correct answer: a**rch** of right foot  **Score:**  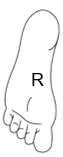  Heel (1)  Arch (2)  Ball (1)  Ahead of toes – not touching (0)  **Notes:** |
| 3.Keep the left foot in place, and move **the R foot** back and forth to then land so the heel of the foot to hoover over the pen.  If I were to set your foot down, which part of your foot would touch the pen?  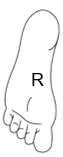 | 3. Correct answer: **heel** of right foot (even)  **Score:**  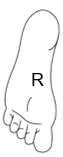  Heel (2)  Arch (1)  Ball (0)  Ahead of toes – not touching (0)  **Notes:** |
| 4.Keep the left foot in place, and move **the R foot** back and forth to then land so the balls of the foot to hoover over the pen.  If I were to set your foot down, which part of your foot would touch the pen?  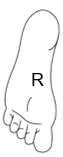 | 4. Correct answer: **arch** of right foot  **Score:**  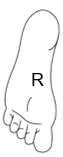  Heel (1)  Arch (2)  Ball (1)  Ahead of toes – not touching (0)  **Noes:** |
| 5.Keep the right foot in place, and move **the L foot** back and forth to then land so the balls of the foot to hoover over the pen.  If I were to set your foot down, which part of your foot would touch the pen?  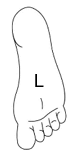 | 5. Correct answer: **ball** of left foot  **Score:**  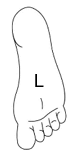  Heel (0)  Arch (1)  Ball (2)  Ahead of toes – not touching (1)  **Notes:** |
| 6.Keep the right foot in place, and move **the L foot** back and forth to then land so the balls of the foot to hoover over the pen.  If I were to set your foot down, which part of your foot would touch the pen?  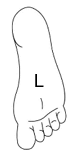 | 6. Correct answer: **toes** of left foot  **Score:**  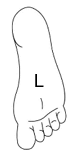  Heel (0)  Arch (1)  Ball (1)  Ahead of toes – not touching (2)  **Notes:** |
| 7.Keep the right foot in place, and move **the L foot** back and forth to then land so the balls of the foot to hoover over the pen.  If I were to set your foot down, which part of your foot would touch the pen?  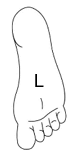 | 7. Correct answer: **heel** of left foot  **Score:**  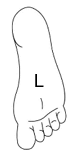  Heel (2)  Arch (1)  Ball (0)  Ahead of toes – not touching (0)  **Notes:** |
| 8.Keep the right foot in place, and move **the L foot** back and forth to then land so the balls of the foot to hoover over the pen.  If I were to set your foot down, which part of your foot would touch the pen?  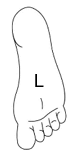 | 8. Correct answer: **ball** of left foot  **Score:**    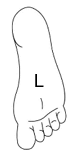  Heel (0)  Arch (1)  Ball (2)  Ahead of toes – not touching (1)  **Notes:** |

Domain 2: Lower limbs has a total score of 32. HIGHER score is BETTER. (i.e., less deficit in MBR)

**UPPER LIMBS**

**Preparation:**

- The patient should sit on the edge of the treatment table with feet flat on the floor and legs at least 90-degrees.
- If the patient is unable to reach the ground, place a box under the feet to ensure the correct positioning.

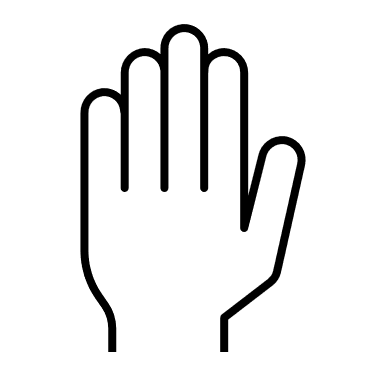

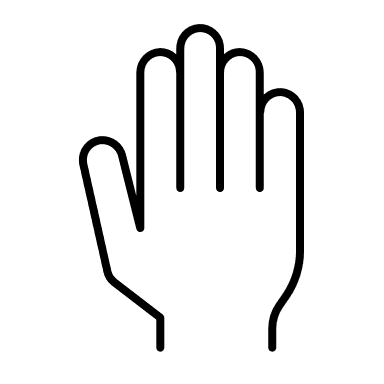

~2f

- The patient should have their arms on a table with hands flat and elbows off of the edge. Arms should be a little less than 90-degrees.
- Stand on the side of the patient, not in front.
- Place an object on the table about 2 feet away

**2d**. We will now go through the same exercise with your upper body. I will move your hands and then ask: Are your hands at the same distance from the object or is one closer? Please close your eyes and I will begin.

| **Movement** | **Score** |
| --- | --- |
| 1. 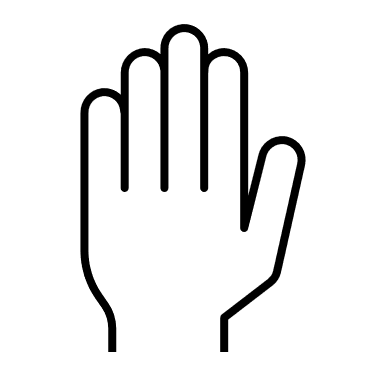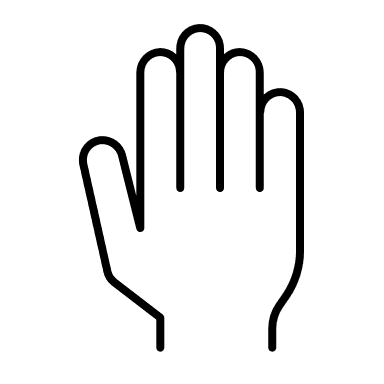 L  R  R  L  **Left** hand moves and lands behind right hand 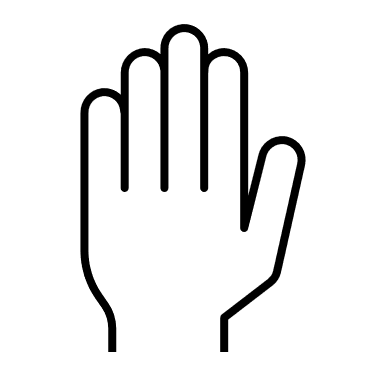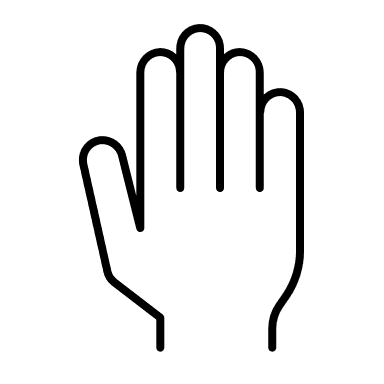 R  L 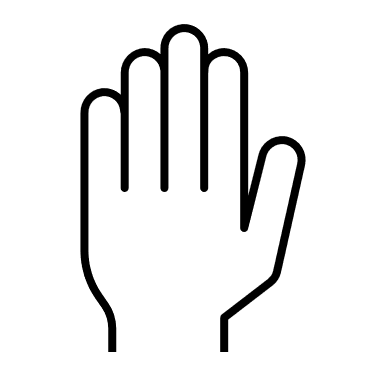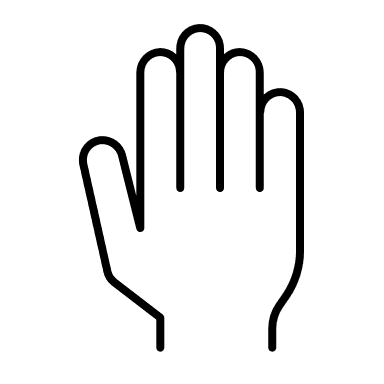 | 1.Correct answer: R is closer  **Score:**  Correct (1)  Incorrect (0)  **Notes:** |
| 2.  **Left** hand moves and lands in front of right hand 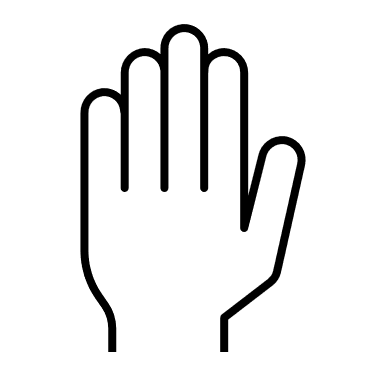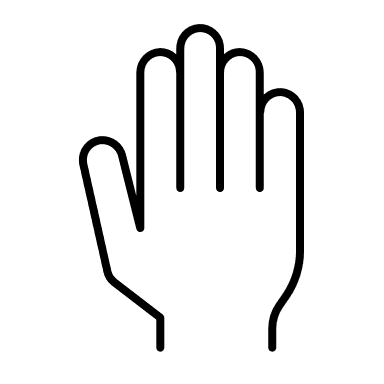 L  R | 2. Correct answer: L is closer  **Score:**  Correct (1)  Incorrect (0)  **Notes:** |
| 3.  **Left** hand moves and lands behind right hand 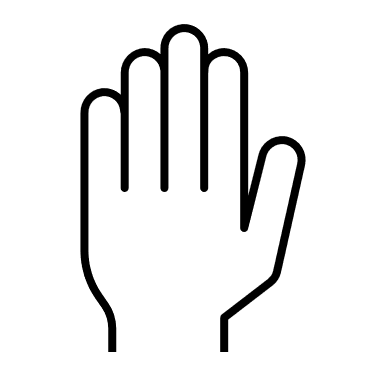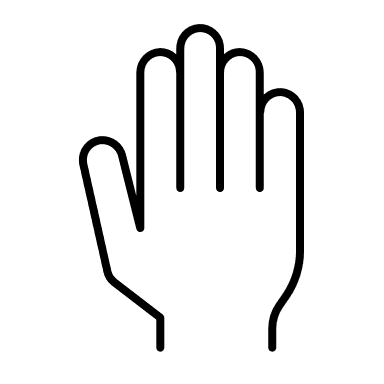 L  R 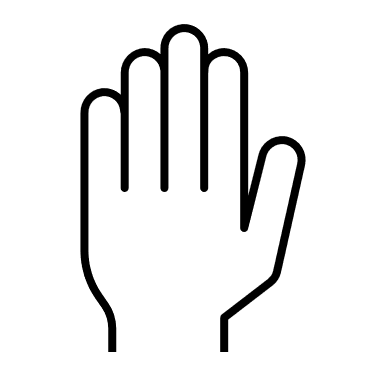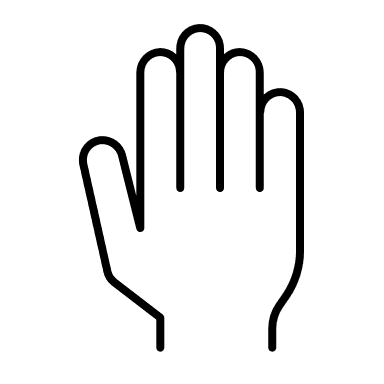 L  R | 3. Correct answer: R is closer  **Score:**  Correct (1)  Incorrect (0)  **Notes:** |
| 4.  **Left** hand moves and lands even with right hand 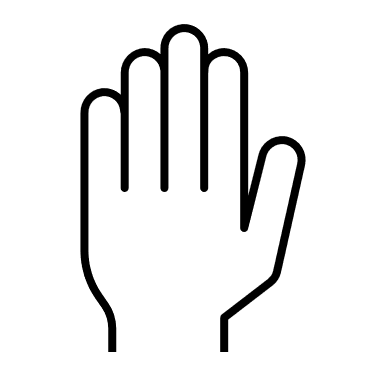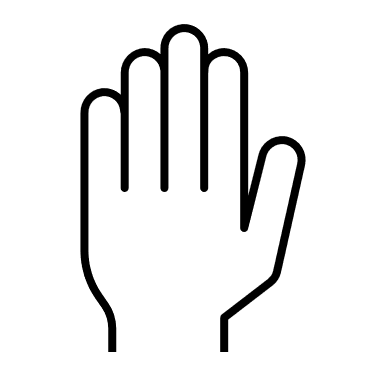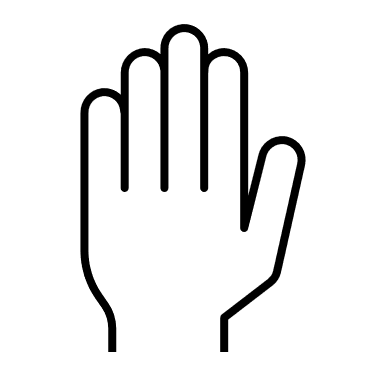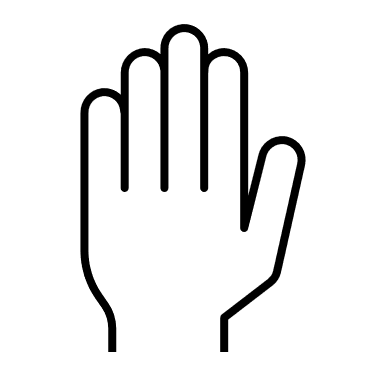 L  R  R  L | 4. Correct answer: Even  **Score:**  Correct (1)  Incorrect (0)  **Notes:** |
| 5. *Instructions:* Please re-center before moving to the R hand.  **Right** hand moves and lands in front left hand  L 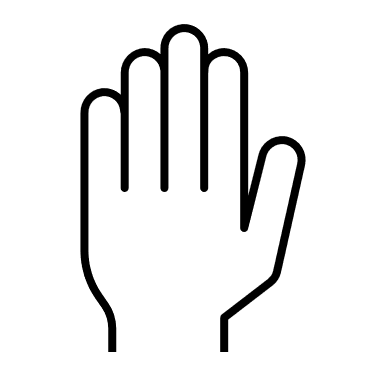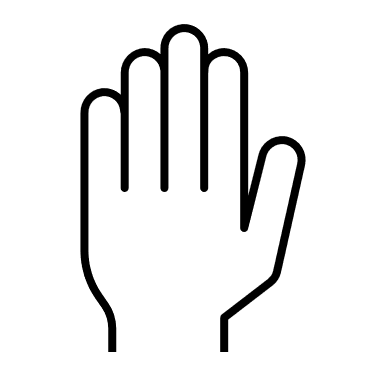 R  R  L 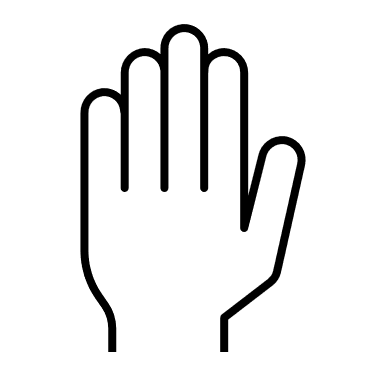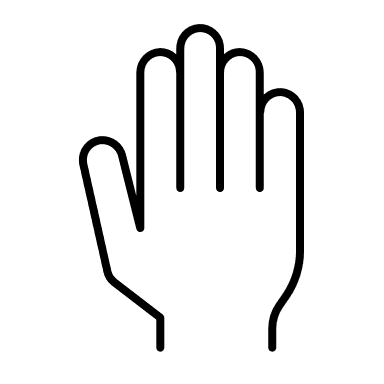 | 5. Correct answer: R is closer  **Score:**  Correct (1)  Incorrect (0)  **Notes:** |
| 6.  **Right** hand moves and lands even with left hand  L  R  R  L 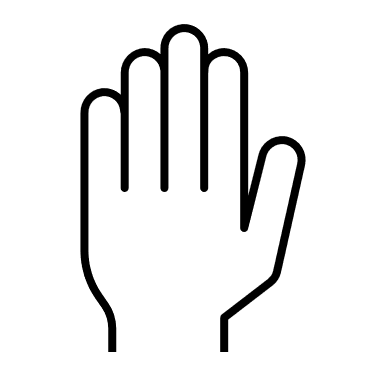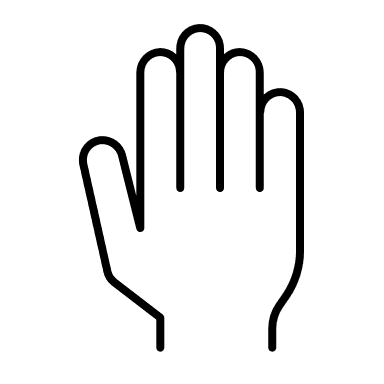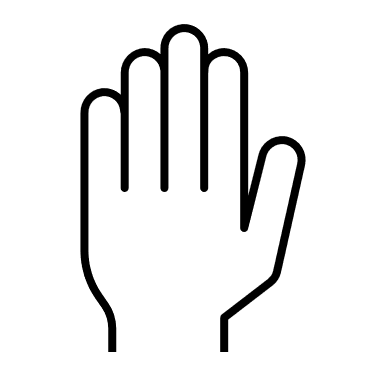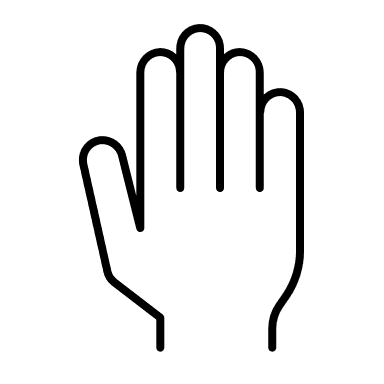 | 6. Correct answer: Even  **Score:**  Correct (1)  Incorrect (0)  **Notes:** |
| 7.  **Right** hand moves and lands behind left hand  L  R  L  R | 7. Correct answer: L is closer  **Score:**  Correct (1)  Incorrect (0)  **Notes:** |
| 8.  **Right** hand moves and lands in front left hand  R  L  R  L | 8. **Correct answer:** R is closer  **Score:**  Correct (1)  Incorrect (0)  **Notes:** |

**2e.** I will move one of your hands and ask which part of one hand your fingertips align with. Please look at the handout for a visual of your options. The fingertips of one hand with align with either your fingers (even), your knuckles, or your wrist.

| **Movement** | **Score** |
| --- | --- |
| 1.Move the **L hand** back and forth to then land at knuckles of right hand.  The tip of your left hand, is in line with which part of your right hand?  L  R | 1. Correct answer: **knuckles** of right hand.  **Score:**  Correct (1)  Incorrect (0)  **Notes:** |
| 2.Without re-centering, move the **L hand** back  and forth to then land at wrist of right hand.  The tip of your left hand, is in line with which part of your right hand?  L  R | 2. Correct answer: **wrist** of right hand.  **Score:**  Correct (1)  Incorrect (0)  **Notes:** |
| 3.Without re-centering, move the **L hand** back  and forth to then land at the knuckles of the right hand.  The tip of your left hand, is in line with which part of your right hand?  R  L | 3. Correct answer: **knuckles** of right hand.  **Score:**  Correct (1)  Incorrect (0)  **Notes:** |
| 4.Without re-centering, move the **L hand** back  and forth to then land at the wrist of right hand.  The tip of your left hand, is in line with which part of your right hand?  L  R | 4. Correct answer: **wrist** of right hand  **Score:**  Correct (1)  Incorrect (0)  **Notes:** |
| 5.*Instructions:* you may re-center before moving to the R hand.  Move the **R hand** back and forth to then land at fingertips/even of the left hand.  The tip of your left hand, is in line with which part of your right hand?  R  L | 5. Correct answer: even  **Score:**  Correct (1)  Incorrect (0)  **Notes:** |
| 6.Without re-centering, move the **R hand** back  and forth to then land at the knuckles of the left hand.  The tip of your left hand, is in line with which part of your right hand?  R  L | 6. Correct answer: **knuckles** of left hand.  **Score:**  Correct (1)  Incorrect (0)  **Notes: wrist** |
| 7.Without re-centering, move the **R hand** back  and forth to then land at the wrist of the left hand.  The tip of your left hand, is in line with which part of your right hand?  R  L | 7. Correct answer: **wrist** of the left hand.  **Score:**  Correct (1)  Incorrect (0)  **Notes:** |
| 8.Without re-centering, move the **R hand** back  and forth to then land at the wrist of the left hand.  The tip of your left hand, is in line with which part of your right hand?  L  R | 8. Correct answer: **wrist** of the left hand  **Score:**  Correct (1)  Incorrect (0)  **Notes:** |

**2f.** Now I will lift up your hand and place a pen underneath the palm. The pen will remain in the same position, but I will move your hand in the air above it. For each movement, I will ask, If I were to set your hand down, which part of your hand would touch the pen. Please use this hand-out for reference.

The options will be in front of fingers (not touching), palm, and wrist.

| **Movement** | **Score** |
| --- | --- |
| 1.If I were to set your hand down, which part of your hand would touch the pen  R | 1. Correct answer: **not touching**  **Score:**  Correct (1)  Incorrect (0)  **Notes:** |
| 2.If I were to set your hand down, which part of your hand would touch the pen    R | 2. Correct answer: **palm**  **Score:**  Correct (1)  Incorrect (0)  **Notes:** |
| 3.If I were to set your hand down, which part of your hand would touch the pen  R | 3. Correct answer: **not touching**  **Score:**  Correct (1)  Incorrect (0)  **Notes:** |
| 4.If I were to set your hand down, which part of your hand would touch the pen    R | 4. Correct answer: **wrist**  **Score:**  Correct (1)  Incorrect (0)  **Notes:** |
| 5.If I were to set your hand down, which part of your hand would touch the pen  L | 5. Correct answer: **palm**  **Score:**  Correct (1)  Incorrect (0)  **Notes:** |
| 6.If I were to set your hand down, which part of your hand would touch the pen    L | 6. Correct answer: **wrist**  **Score:**  Correct (1)  Incorrect (0)  **Notes:** |
| 7.If I were to set your hand down, which part of your hand would touch the pen    L | 7. Correct answer: **palm**  **Score:**  Correct (1)  Incorrect (0)  **Notes:** |
| 8.If I were to set your hand down, which part of your hand would touch the pen  L | 8. Correct answer: **not touching**  **Score:**  Correct (1)  Incorrect (0)  **Notes:** |

Domain 2: upper limbs, has a total score of 24. HIGHER score is BETTER. (i.e., less deficit in MBR)

**Domain 3: SPATIAL AWARENESS**

**Preparation:**

- The patient should sit on the edge of the treatment table with feet flat on the floor and legs at least 90-degree angle.
- If the patient is unable to reach the ground, place a box under the feet to ensure the correct positioning.
- The patient can have shoes on or off for this task.
- Place yourself with your hand only 2 feet away from the body part.

**3a.** The goal of this task is to evaluate body awareness in space. I will have you close your eyes and I will take your hand in mine and make it a pointed finger. I will guide your finger to certain positions and will ask *“*In this position, if I were to keep moving your palm in this direction would it touch your body? If yes, which part?”

*Instructions:* **use right hand**

| **Movement** | **Score** |
| --- | --- |
| 1.Take palm and point it in between the two knees, **(missing).** | 1.Correct answer: **missing body, in between both knees**  **Score:**  Correct (1)  Incorrect (0)  **Patient response:** |
| 2.Take palm and point it towards **right upper leg.** | 2. Correct answer: **on right upper leg**  **Score:**  Correct (1)  Incorrect (0)  **Patient response:** |
| 3.Take palm and point it outside of the right knee cap, **(missing)** | 3.Correct answer: **missing, too far to the right of the right knee**  **Score:**  Correct (1)  Incorrect (0)  **Patient response:** |
| 4.Take palm and point it towards right knee, but too far forward **(missing)** | 4.Correct answer: not touching, too far forward past knee.  **Score:**  Correct (1)  Incorrect (0)  **Patient response:** |
| 5.Take palm and point it towards **left upper leg.** | 5.Correct answer: **left upper leg/thigh**  **Score:**  Correct (1)  Incorrect (0)  **Patient response:** |
| 6.Take palm and point it towards **left knee** cap. | 6.Correct answer: **left knee**  **Score:**  Correct (1)  Incorrect (0)  **Patient response:** |
| 7.Take palm and point it towards the space too far to the left of the left leg. **(missing)** | 7.Correct answer: **no, too far to the outside of the upper left leg.**  **Score:**  Correct (1)  Incorrect (0)  **Patient response:** |
| 8.Take palm and point it towards the **right knee** (touching). | 8.Correct answer: **right** **knee**  **Score:**  Correct (1)  Incorrect (0)  **Patient response:** |

Domain 3 has a total score of 8. HIGHER score is BETTER. (i.e., less deficit in MBR)

**Domain 4: BODY LOCALIZATION TASK**

**Preparation:**

- The patient should sit in either a chair with arm rests or in a wheel chair.
- The chair should be positioned in the center of the room.
- Ask the patient if they have any hearing issues and record if they do.

**4a.** For this task, I will make a snapping noise at different locations around your body at about a 2 ft. distance. Once the task starts, I will ask you to close your eyes and answer each question as it comes. For each snap, I will ask you if the snapping noise was in the front of your body or behind your body. Then, I will ask whether the snapping sound was at your head level or feet level. Please look at the handout to get a visual of the options. Please close your eyes, and I will begin.

*Instructions:* Place yourself standing on either the left or right side of the patient for numbers 1-8. Try your best not to make any sounds while placing your hand as far to the front center or back center of the body before making the snap.

| **Movement** | **Score** |
| --- | --- |
| 1 | **Score front to back:**  Front (1)  Back (0)  **Score level:**  Head (1)  Feet (0) |
| 2 | **Score front to back:**  Front (1)  Back (0)  **Score level:**  Feet (1)  Head (0) |
| 3 | **Score front to back:**  Back (1)  Front (0)  **Score level:**  Feet (1)  Head (0) |
| 4 | **Score front to back:**  Front (1)  Back (0)  **Score level:**  Head (1)  Feet (0) |
| 5 | **Score front to back:**  Back (1)  Front (0)  **Score level:**  Head (1)  Feet (0) |
| 6 | **Score front to back:**  Back (1)  Front (0)  **Score level:**  Feet (1)  Head (0) |
| 7 | **Score front to back:**  Front (1)  Back (0)  **Score level:**  Feet (1)  Head (0) |
| *8* | **Score front to back:**  Front (1)  Back (0)  **Score level:**  Feet (1)  Head (0) |

**4b.** Now, for each snap, I will stay only on your dominant side and will ask you if the snapping noise was at your head level, hip level, or feet level.

| **Movement** | **Score** |
| --- | --- |
|  | **Score:**  Head (1)  Hip (0)  Feet (0) |
|  | **Score:**  Feet (1)  Hip (0)  Head (0) |
|  | **Score:**  Feet (1)  Hip (0)  Head (0) |
|  | **Score:**  Head (1)  Hip (0)  Feet (0) |
|  | **Score:**  Hip (1)  Head (0)  Feet (0) |
|  | **Score:**  Head (1)  Hip (0)  Feet (0) |
|  | **Score:**  Hip (1)  Head (0)  Feet (0) |
|  | **Score:**  Feet (1)  Head (0)  Hip (0) |

Domain 4 has a total score of 16. HIGHER score is BETTER. (i.e., less deficit in	MBR).

**SCI-BodyMap Self Report Section**

**SCORING**

Instructions for PT/OT: For adults with SCI who are unable or who prefer not to, you may shade in the body part for them. Although these are self-reports, it is highly encouraged that a PT/OT explains the items thoroughly to make sure it is correctly interpreted by the participant.

Scoring: **Domain 5: Body Awareness and Domain 6: Sensation** – HIGHER is BETTER (i.e., less deficit in MBR) For each of the 20 body regions listed in Domain 7, there will be a score of:

**(**3) Uncolored

(2) Only Yellow

(1) Yellow + Blue

(0) Only Blue

Sum of all body regions__/60

Scoring: **Domains 7: Item a** – LOWER is BETTER (i.e., less Neuropathic pain) For each of the 20 body regions listed in Domain 7, there will be a score of:

(0) Uncolored
(1) Only Yellow
(2) Only Blue
(3) Yellow + Blue

Sum of all body regions__/60

Scoring: **Domains 7, Items 7a – 7i** – LOWER is BETTER (i.e., less Neuropathic pain). For each of the 20 body regions listed in Domain 7, take the sum of each item. Please interpret each item separately. There will be a total score for Yellow and a total score for Blue for each item (see below).

Frequency Blue ___ /80

Duration Blue ___ /80

Pain most often Blue ___ /80

Highest pain Blue___ /80

Frequency Yellow ___ /80

Duration Yellow ___ /80

Pain most Yellow ___ /80

Highest pain Yellow ___ /80

**THERE WILL BE ONE CHART FOR YELLOW AND ONE CHART FOR BLUE IF THE PARTICIPANT HAS BOTH REPORTED PAIN AREAS**

| **Frequency:** How often do you experience neuropathic pain in a typical week? | **Duration:** How long did each instance last? (If they varied, what was the most common duration) | **Intensity:** How would you describe the intensity of this feeling MOST of the time in a typical week? | **Intensity:** How would you describe the HIGEST  intensity of this feeling in a typical week? |
| --- | --- | --- | --- |
| 0)Had no neuropathic pain  1)Less than once a day   2) 1-5 times a day 3) 5 or more times a day 4) 100 percent of the time | 0)Had no neuropathic pain  1)Less than once a day   2) 1-5 times a day 3) 5 or more times a day 4) 100 percent of the time | 0)Had no neuropathic pain  1)Light  2)Moderate  3)Severe  4)Excruciating | 0)Had no neuropathic pain  1)Light  2)Moderate  3)Severe  4)Excruciating |

**Domain 7: NEUROPATHIC PAIN (items 7b-7i)**

For each body region, please answer the following questions (note: if you left the body region uncolored, then the answer should be “had no neuropathic pain”)

***Fill in one for Yellow and one for Blue (if needed)**

| **Part of the body** | **7b/f. Frequency:** How often do you experience neuropathic pain in a typical week? | **7c/g. Duration:** How long did each instance last? (If they varied, what was the most common duration) | **7d/h. Intensity:** How would you describe the intensity of this feeling MOST of the time in a typical week? | **7e/i. Intensity:** How would you describe the HIGEST intensity of this feeling in a typical week? |
| --- | --- | --- | --- | --- |
| 1.**Head** | - Had no neuropathic pain - Less than once a day - 1-5 times a day - 5 or more times a day - 100 percent of the time | - Had no neuropathic pain - Less than 1 hour - 1-5 hours - Over 5 hours-almost always - 100 percent of the time | - Had no neuropathic pain - Light - Moderate - Severe - Excruciating | - Had no neuropathic pain - Light - Moderate - Severe - Excruciating |
| 2.**Throat/Neck** | - Had no neuropathic pain - Less than once a day - 1-5 times a day - 5 or more times a day - 100 percent of the time | - Had no neuropathic pain - Less than 1 hour - 1-5 hours - Over 5 hours-almost always - 100 percent of the time | - Had no neuropathic pain - Light - Moderate - Severe - Excruciating | - Had no neuropathic pain - Light - Moderate - Severe - Excruciating |
| 3.**Chest** | - Had no neuropathic pain - Less than once a day - 1-5 times a day - 5 or more times a day - 100 percent of the time | - Had no neuropathic pain - Less than 1 hour - 1-5 hours - Over 5 hours-almost always - 100 percent of the time | - Had no neuropathic pain - Light - Moderate - Severe - Excruciating | - Had no neuropathic pain - Light - Moderate - Severe - Excruciating |
| 4.**Upper back** | - Had no neuropathic pain - Less than once a day - 1-5 times a day - 5 or more times a day - 100 percent of the time | - Had no neuropathic pain - Less than 1 hour - 1-5 hours - Over 5 hours-almost always - 100 percent of the time | - Had no neuropathic pain - Light - Moderate - Severe - Excruciating | - Had no neuropathic pain - Light - Moderate - Severe - Excruciating |
| 5.**Belly** | - Had no neuropathic pain - Less than once a day - 1-5 times a day - 5 or more times a day - 100 percent of the time | - Had no neuropathic pain - Less than 1 hour - 1-5 hours - Over 5 hours-almost always - 100 percent of the time | - Had no neuropathic pain - Light - Moderate - Severe - Excruciating | - Had no neuropathic pain - Light - Moderate - Severe - Excruciating |
| 6.**Mid/Lower back** | - Had no neuropathic pain - Less than once a day - 1-5 times a day - 5 or more times a day - 100 percent of the time | - Had no neuropathic pain - Less than 1 hour - 1-5 hours - Over 5 hours-almost always - 100 percent of the time | - Had no neuropathic pain - Light - Moderate - Severe - Excruciating | - Had no neuropathic pain - Light - Moderate - Severe - Excruciating |
| 7.**Right shoulder/upper arm** | - Had no neuropathic pain - Less than once a day - 1-5 times a day - 5 or more times a day - 100 percent of the time | - Had no neuropathic pain - Less than 1 hour - 1-5 hours - Over 5 hours-almost always - 100 percent of the time | - Had no neuropathic pain - Light - Moderate - Severe - Excruciating | - Had no neuropathic pain - Light - Moderate - Severe - Excruciating |
| 8.**Left shoulder/upper arm** | - Had no neuropathic pain - Less than once a day - 1-5 times a day - 5 or more times a day - 100 percent of the time | - Had no neuropathic pain - Less than 1 hour - 1-5 hours - Over 5 hours-almost always - 100 percent of the time | - Had no neuropathic pain - Light - Moderate - Severe - Excruciating | - Had no neuropathic pain - Light - Moderate - Severe - Excruciating |
| 9.**Right elbow/forearm** | - Had no neuropathic pain - Less than once a day - 1-5 times a day - 5 or more times a day - 100 percent of the time | - Had no neuropathic pain - Less than 1 hour - 1-5 hours - Over 5 hours-almost always - 100 percent of the time | - Had no neuropathic pain - Light - Moderate - Severe - Excruciating | - Had no neuropathic pain - Light - Moderate - Severe - Excruciating |
| 10.**Left elbow/forearm** | - Had no neuropathic pain - Less than once a day - 1-5 times a day - 5 or more times a day - 100 percent of the time | - Had no neuropathic pain - Less than 1 hour - 1-5 hours - Over 5 hours-almost always - 100 percent of the time | - Had no neuropathic pain - Light - Moderate - Severe - Excruciating | - Had no neuropathic pain - Light - Moderate - Severe - Excruciating |
| 11.**Right wrist/hand** | - Had no neuropathic pain - Less than once a day - 1-5 times a day - 5 or more times a day - 100 percent of the time | - Had no neuropathic pain - Less than 1 hour - 1-5 hours - Over 5 hours-almost always - 100 percent of the time | - Had no neuropathic pain - Light - Moderate - Severe - Excruciating | - Had no neuropathic pain - Light - Moderate - Severe - Excruciating |
| 12.**Left wrist/hand** | - Had no neuropathic pain - Less than once a day - 1-5 times a day - 5 or more times a day - 100 percent of the time | - Had no neuropathic pain - Less than 1 hour - 1-5 hours - Over 5 hours-almost always - 100 percent of the time | - Had no neuropathic pain - Light - Moderate - Severe - Excruciating | - Had no neuropathic pain - Light - Moderate - Severe - Excruciating |
| 13.**Pelvis/Groin** | - Had no neuropathic pain - Less than once a day - 1-5 times a day - 5 or more times a day - 100 percent of the time | - Had no neuropathic pain - Less than 1 hour - 1-5 hours - Over 5 hours-almost always - 100 percent of the time | - Had no neuropathic pain - Light - Moderate - Severe - Excruciating | - Had no neuropathic pain - Light - Moderate - Severe - Excruciating |
| 14.**Buttocks** | - Had no neuropathic pain - Less than once a day - 1-5 times a day - 5 or more times a day - 100 percent of the time | - Had no neuropathic pain - Less than 1 hour - 1-5 hours - Over 5 hours-almost always - 100 percent of the time | - Had no neuropathic pain - Light - Moderate - Severe - Excruciating | - Had no neuropathic pain - Light - Moderate - Severe - Excruciating |
| 15.**Right hip/upper leg** | - Had no neuropathic pain - Less than once a day - 1-5 times a day - 5 or more times a day - 100 percent of the time | - Had no neuropathic pain - Less than 1 hour - 1-5 hours - Over 5 hours-almost always - 100 percent of the time | - Had no neuropathic pain - Light - Moderate - Severe - Excruciating | - Had no neuropathic pain - Light - Moderate - Severe - Excruciating |
| 16.**Left hip/upper leg** | - Had no neuropathic pain - Less than once a day - 1-5 times a day - 5 or more times a day - 100 percent of the time | - Had no neuropathic pain - Less than 1 hour - 1-5 hours - Over 5 hours-almost always - 100 percent of the time | - Had no neuropathic pain - Light - Moderate - Severe - Excruciating | - Had no neuropathic pain - Light - Moderate - Severe - Excruciating |
| 17.**Right knee/lower leg** | - Had no neuropathic pain - Less than once a day - 1-5 times a day - 5 or more times a day - 100 percent of the time | - Had no neuropathic pain - Less than 1 hour - 1-5 hours - Over 5 hours-almost always - 100 percent of the time | - Had no neuropathic pain - Light - Moderate - Severe - Excruciating | - Had no neuropathic pain - Light - Moderate - Severe - Excruciating |
| 18.**Left knee/lower leg** | - Had no neuropathic pain - Less than once a day - 1-5 times a day - 5 or more times a day - 100 percent of the time | - Had no neuropathic pain - Less than 1 hour - 1-5 hours - Over 5 hours-almost always - 100 percent of the time | - Had no neuropathic pain - Light - Moderate - Severe - Excruciating | - Had no neuropathic pain - Light - Moderate - Severe - Excruciating |
| 19.**Right ankle/foot** | - Had no neuropathic pain - Less than once a day - 1-5 times a day - 5 or more times a day - 100 percent of the time | - Had no neuropathic pain - Less than 1 hour - 1-5 hours - Over 5 hours-almost always - 100 percent of the time | - Had no neuropathic pain - Light - Moderate - Severe - Excruciating | - Had no neuropathic pain - Light - Moderate - Severe - Excruciating |
| 20.**Left ankle/foot** | - Had no neuropathic pain - Less than once a day - 1-5 times a day - 5 or more times a day - 100 percent of the time | - Had no neuropathic pain - Less than 1 hour - 1-5 hours - Over 5 hours-almost always - 100 percent of the time | - Had no neuropathic pain - Light - Moderate - Severe - Excruciating | - Had no neuropathic pain - Light - Moderate - Severe - Excruciating |
